# Longitudinal characterization of *Plasmodium* inter-species interactions during a period of increasing prevalence of *Plasmodium ovale*

**DOI:** 10.1101/2019.12.28.19015941

**Authors:** Hoseah M. Akala, Oliver Watson, Kenneth K. Mitei, Dennis W. Juma, Robert Verity, Luiser A. Ingasia, Benjamin H. Opot, Raphael O. Okoth, Gladys C. Chemwor, Jackline A. Juma, Edwin W. Mwakio, Nicholas Brazeau, Agnes C. Cheruiyot, Redemptah A. Yeda, Maureen N. Maraka, Charles O. Okello, David P. Kateete, Jim Ray Managbanag, Ben Andagalu, Bernhards R. Ogutu, Edwin Kamau

## Abstract

**Background:** The epidemiology and severity of non-falciparum malaria in endemic settings has garnered limited attention. We aimed to characterize the prevalence, interaction, clinical risk factors and temporal trends of non-falciparum malaria in endemic settings of Kenya.

**Methods:** We diagnosed and analyzed infecting malaria species via PCR in 2027 clinical samples collected between 2008 and 2016. Descriptive statistics were used to describe the prevalence and distribution of *Plasmodium* species. A statistical model was designed and used for estimating the frequency of *Plasmodium* species and assessing inter-species interactions. Mixed effect linear regression models with random intercepts for each location was used to test for change in prevalence over time.

**Findings:** 72•5% of the samples were *P. falciparum* single species infections, 25·8% were mixed infections and only 1•7% occurred as single non-falciparum species infections. 23•1% were mixed infections containing *P. ovale*. A likelihood-based model calculation of the population frequency of each species estimated a significant within-host interference between *P. falciparum* and *P. ovale curtisi*. Mixed-effect logistic regression models identified a significant increase of *P. ovale wallikeri* and *P. ovale curtisi* species over time with reciprocal decrease in *P. falciparum single species* and *P. malariae*. The risk of *P. falciparum* infections presenting with fever was 0•43 times less likely if co-infected with *P. malariae*.

**Interpretation:** Findings show higher prevalence of non-falciparum malaria than expected. The proportion of infections that were positive for infection by *P. ovale wallikeri* and *P. ovale curtisi* was observed to significantly increase over the period of study which could be due to attenuated responsiveness to malaria drug treatment on these species. The increase in frequency of *P. ovale spp* in Kenya could threaten malaria control effort in Kenya and pose increased risk of malaria to travelers.

**Funding:** AFHSB and its GEIS Section

## Introduction

Malaria control programmes in sub-Saharan Africa (sSA) have mostly focused on *Plasmodium falciparum*, the predominant cause of lethal malaria. Although widely distributed in malaria-endemic regions including sSA,^1-3^ information on the epidemiology of non-falciparum malaria caused by *P. ovale* spp. and *P. malariae* is scarce. Further, the severity of the disease caused by these species, which is thought to be milder compared to *P. falciparum*, is not well studied in the endemic human populations.^3^ Most of the clinical data for non-falciparum malaria has been obtained mostly from travelers returning from malaria endemic areas.^3, 4^

*P. ovale* spp. and *P. malariae* often occur in complex coinfections with *P. falciparum* which complicates the ability to accurately detect these infections using light microscopy^1^ or malaria rapid diagnostic tests (mRDT).^5^ This is further exacerbated by the fact that these infections often occur at low parasitemia,^6^ which likely contributes to the underestimation on the prevalence of non-falciparum in sSA. However, molecular and serological studies have been shown to be more sensitive, revealing that non-falciparum infections are more prevalent than previously estimated,^7, 8^ with one study measuring seroprevalence of *P. ovale* spp. and *P. malariae* at 57% and 45% respectively, in asymptomatic populations in Benin.^8^

Artemisinin-combination therapies (ACTs) are the first-line treatment for uncomplicated *P. falciparum* malaria infections in most malaria endemic countries. However, there is limited *in vivo* data available on the efficacy of these drugs against non-falciparum malaria.^7^ Further, most therapeutic efficacy studies exclude mixed infections as part of their enrolment criteria. These studies mostly use microscopy to assess drug efficacy, which is less sensitive compared to PCR.^9^ Recent studies have detected persistent non-falciparum parasites after treatment with ACTs when assessed by PCR.^7, 10, 11^ There is thus a need to characterize species specific responses to ACTs, which can help inform the efficacy of ACT regimens in treating non-falciparum infections in malaria endemic regions. This is of importance both in efforts to control malaria in endemic regions and for imported malaria cases as it has been shown non-falciparum contribute to a large proportion of imported uncomplicated and complicated malaria in non-endemic countries.^4, 12, 13^

Molecular surveillance provides more accurate speciation data as recently demonstrated by a cross-sectional study in western Kenya where the overall prevalence of *Plasmodium* spp. was estimated to be 37•1% (13•2% non-falciparum malaria) by PCR versus 19•9% (1•6% non-falciparum malaria) by microscopy.^14^ To further our understanding on the spatial and temporal trends in non-falciparum malaria, we conducted a longitudinal study in four different malaria endemicity zones in Kenya, and collected clinical and malaria speciation data from symptomatic individuals seeking treatment in healthcare facilities. Comprehensive patient data was collected and blood samples were analyzed using highly sensitive speciating real-time PCR (qPCR). We investigated the clinical risk factors associated with non-falciparum infections in a malaria endemic population, and developed and applied a novel statistical framework for exploring whether species occur independently of other species. Lastly, we explored the temporal trends in both single and mixed species infection over the study period across the different endemicity zones.

### Ethics statement

This study was conducted under the approval of the Kenya Medical Research Institute (KEMRI), Scientific and Ethics Review Unit (SERU) and Walter Reed Army Institute of Research (WRAIR) institutional review boards, protocol numbers: KEMRI #1330, WRAIR #1384 entitled “Epidemiology of malaria and drug sensitivity patterns in Kenya.”

## Methods

### Study sites and sampling collection

Samples were collected between 2008 and 2016 from hospitals located in 6 regions that span 4 distinct malaria transmission zones across Kenya (see Figure 1, Table 1)^15^. Consenting patients aged six months and above, presenting at outpatient departments with symptoms of malaria and/or testing positive for uncomplicated malaria by rapid diagnostic test (mRDT; Parascreen® (Pan/Pf), Zephyr Biomedicals, Verna Goa, India) were recruited into the study. Case Report Forms (CSF) were used to collect comprehensive patient information including age, sex, occupation, home of origin, travel history in the last 2-4 weeks, history of malaria infection and treatment, chief complaint, other complaints (such as headache, vomiting, coughing, diarrhea), body temperature and body weight. 2-3 ml of whole blood was collected for mRDT testing and smear preparation. About 100 μl of each sample was spotted on FTA filter paper (Whatman Inc., Bound Brook, New Jersey, USA) for DNA extraction and nucleic acid analysis. The clinician then performed diagnosis by assessing for symptoms such as conjunctival pallor, lymphadenopathy and splenomegaly. Final diagnosis results were based on clinical evaluation confirmed by mRDT and/or microscopy. All malaria positive cases were treated with Coartem® which contains artemether-lumefantrine (AL) in accordance with the Kenya Ministry of Health recommended case management guidelines for uncomplicated malaria.

**Table 1:** Distribution of samples collected across transmission regions

| Region | Hospital | Samples |
| --- | --- | --- |
| Lake endemic zone | Kisumu east sub-county hospital | 887 |
|  | Kombewa sub-county hospital | 437 |
| Coastal endemic zone | Malindi sub-county hospital | 24 |
| Highland epidemic zone | Kericho sub-county hospital | 170 |
|  | Kisii sub-county hospital | 432 |
| Semi-Arid Zone | Marigat sub-county hospital | 77 |

**Figure 1:**
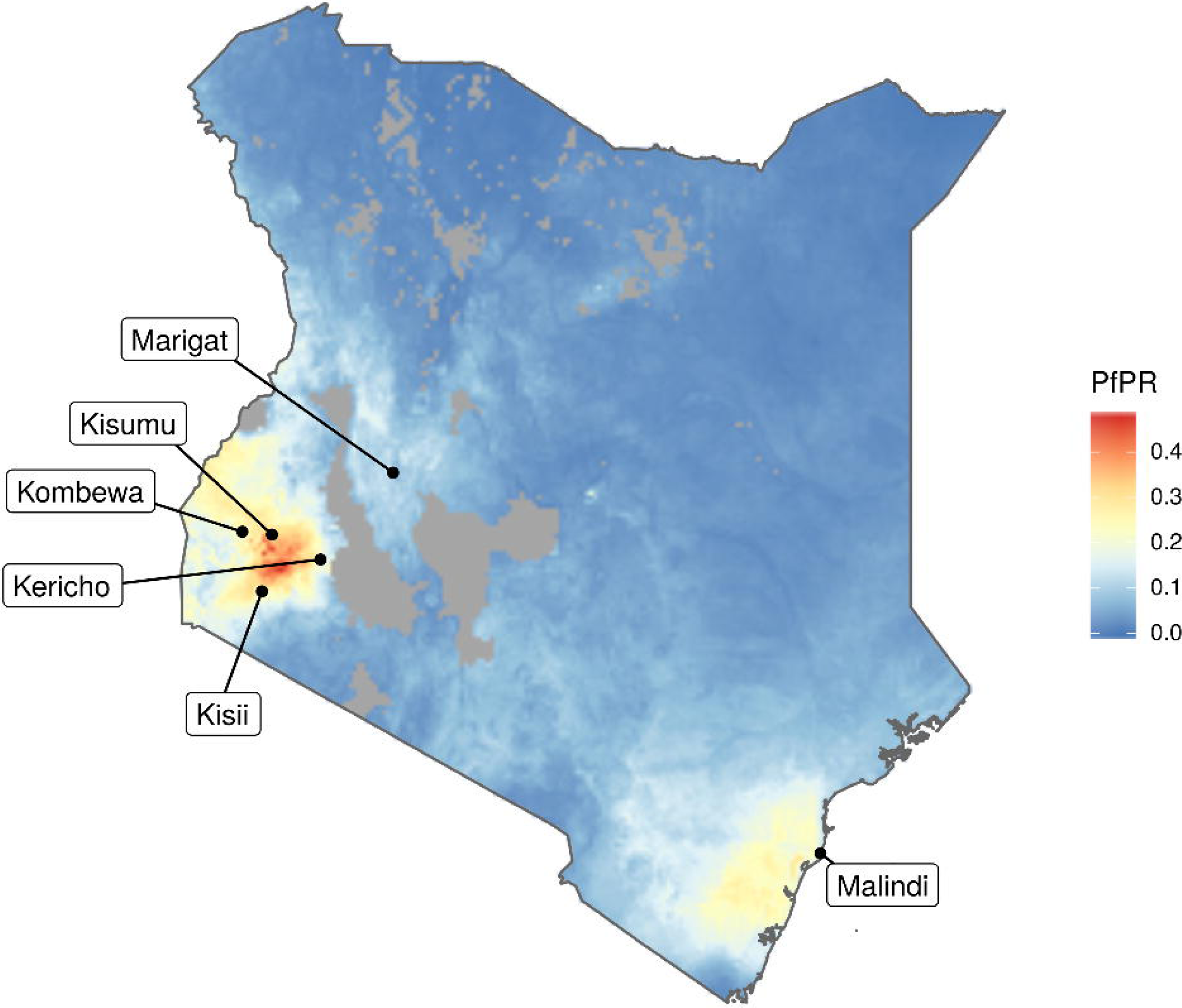
Study sites. Map of Kenya malaria endemicity zones and the six chosen study sites ^26^.

### Genotypic analysis

Genomic DNA was extracted using the QIAamp DNA mini kit (Qiagen, Valencia, CA, USA) as recommended by the manufacturer. The DNA was used for malaria diagnosis using Genus-specific qPCR assay prior to species-specific analyses as previously described for the detection of *Plasmodium* genus and *P. falciparum*.^9^ *For each assay, a negative template control (distilled water) and P. falciparum* DNA Nucleic Acid Tests (NAT)/NIBSC genus-specific positive controls were included to ensure no contamination of the PCR and to validate results. The human housekeeping gene Ribonuclease P (RN*ase*P) was used as an extraction and qPCR assay control.

All samples that were diagnosed as positive for malaria were further characterized for species composition using a separate set of species-specific primers indicated in supplementary Table 1. The assays for characterization of *P. falciparum* and *P. malariae* had identical PCR reaction components and conditions as the Genus-specific qPCR assay except for the primers used. Specifically, FAL R, FAL F, and FAL were used for *P. falciparum* diagnosis while MAL F, MAL R, and MAL PP were used for *P. malariae* diagnosis. Detection of the two *P. ovale* species was conducted using a previously described method.^16, 17^ Reaction components and amplification conditions were identical to the Genus-specific qPCR assay. Each experiment included at least one reaction mixture without DNA as a negative control.

## Statistical analysis

### Estimating the frequency of *Plasmodium* species and assessing inter-species interaction

A statistical model was designed to assess whether there was a significant difference between the observed frequencies of each infection type and the expected frequencies when assuming independent acquisition of different *Plasmodium* species. The approach jointly estimates the population frequency of each *Plasmodium* species and the population mean number of extant infections per individual, and uses these estimates to generate null distributions for the number of single and multi-species infections. The model was extended to statistically assess for the presence of between-species interactions and the best fitting models were identified through comparisons of sample-size corrected Akaike information criterion (AICc).^18^ Full methodology is described in the Supplementary Information.

### Linear modelling of species prevalence and risk factors associated with fever

Summary descriptive statistics were used to describe the prevalence and distribution of *Plasmodium* species and the occurrence of mixed species infections across Kenya between 2008 and 2016. To test for a change in the prevalence of *Plasmodium* species over time, we used a mixed effect linear regression model with random intercepts for each location^19^ using the lme4 R software package.^20^ Due to low numbers of samples collected prior to 2011 in Kombewa, Marigat, and Malindi, samples were grouped according to the 4 endemic zones: highland epidemic (Kericho and Kisii), coastal endemic (Malindi), semi-arid (Marigat), and lake endemic (Kisumu and Kombewa). The samples were also grouped according to the year and month they were collected in, and the regression was weighted according to the number of samples collected in each year and month. This framework was used to assess for changes in the prevalence of infections caused by *P. malariae, P. ovale curtisi, P. ovale wallikeri* as well as the prevalence of infections positive for only *P. falciparum*.

Patient metadata was available for a subset of individuals reporting at clinic, which included the chief symptomatic complaint reported by the patient as well as whether the individual was currently presenting with fever based on temperature measurements on arrival at the hospital. We explored the patterns in the chief complaint with respect to the infecting species composition to assess whether the distribution of infecting species by complaint was predicted by the earlier estimated frequency of *Plasmodium* species. Lastly, we used a mixed effect logistic regression model to assess the risk factors associated with fever presentation at clinic among individuals infected with *P. falciparum*. The included covariates were patient age, sex, time of sample collection, number of malaria attacks in the last year and whether the individual was co-infected with *P. malariae, P. ovale curtisi* or *P. ovale wallikeri*.

## Results

### Sample Collection

During the study period, 3120 study participants from six field sites (sub-county hospitals) were screened, 3058 met inclusion criteria and were enrolled in the study. Of this, patient metadata such as age, sex, chief complaint, body temperature, travel, and malaria infection history, clinical diagnosis, mRDT, and/or microscopy results was collected from 2719 study participants, with 2027 samples successfully analyzed for all *Plasmodium* spp. by speciating qPCR. Table 1 shows number of samples collected from each field site that were qPCR analyzed, grouped per the malaria epidemiological zones, with lake and coastal endemic zones grouped separately. The lake endemic zone accounted for the majority (65•0%) of the samples analyzed whereas the coastal endemic had the least (1•2%).

### *Plasmodium* species composition

*P. falciparum* was the most prevalent species and was present in 98•0% (n = 1986) of the 2027 infections, *P. ovale wallikeri* at 20•1% (n = 405), *P. malariae* at 5•0% (n = 102), and *P. ovale curtisi* at 5•0% (n = 101). 74•2% (n = 1504) of samples carried single parasite species infections while 25•8% (n = 523) had multiple species infections. 0•15% (n = 3) of samples had a mixture of all the four species: *P. falciparum, P. ovale wallikeri, P. ovale curtisi*, and *P. malariae*. The rest of the samples had either two 23•8% (n = 482) or three 1•9% (n = 38) species infections. The most prevalent mixed infections contained *P. falciparum* and *P. ovale wallikeri*. The full composition of the infections observed is shown in Figure 2. When analyzed per study site, Kisumu (urban, lake endemic), and Marigat (semi-arid) had the highest prevalence of *P. falciparum* single infections at 79•4% (95% CI: 76•6% - 81•9%) and 79•2% (95% CI: 68•9% - 86•8%), respectively (Supplementary Figure 1). On the other hand, Kombewa (rural, lake endemic) had the lowest *P. falciparum* single infections at 61•3% (95% CI: 56•7% - 65•8%), which is significantly lower than Kisumu and Marigat (p < 0.01).

**Figure 2:**
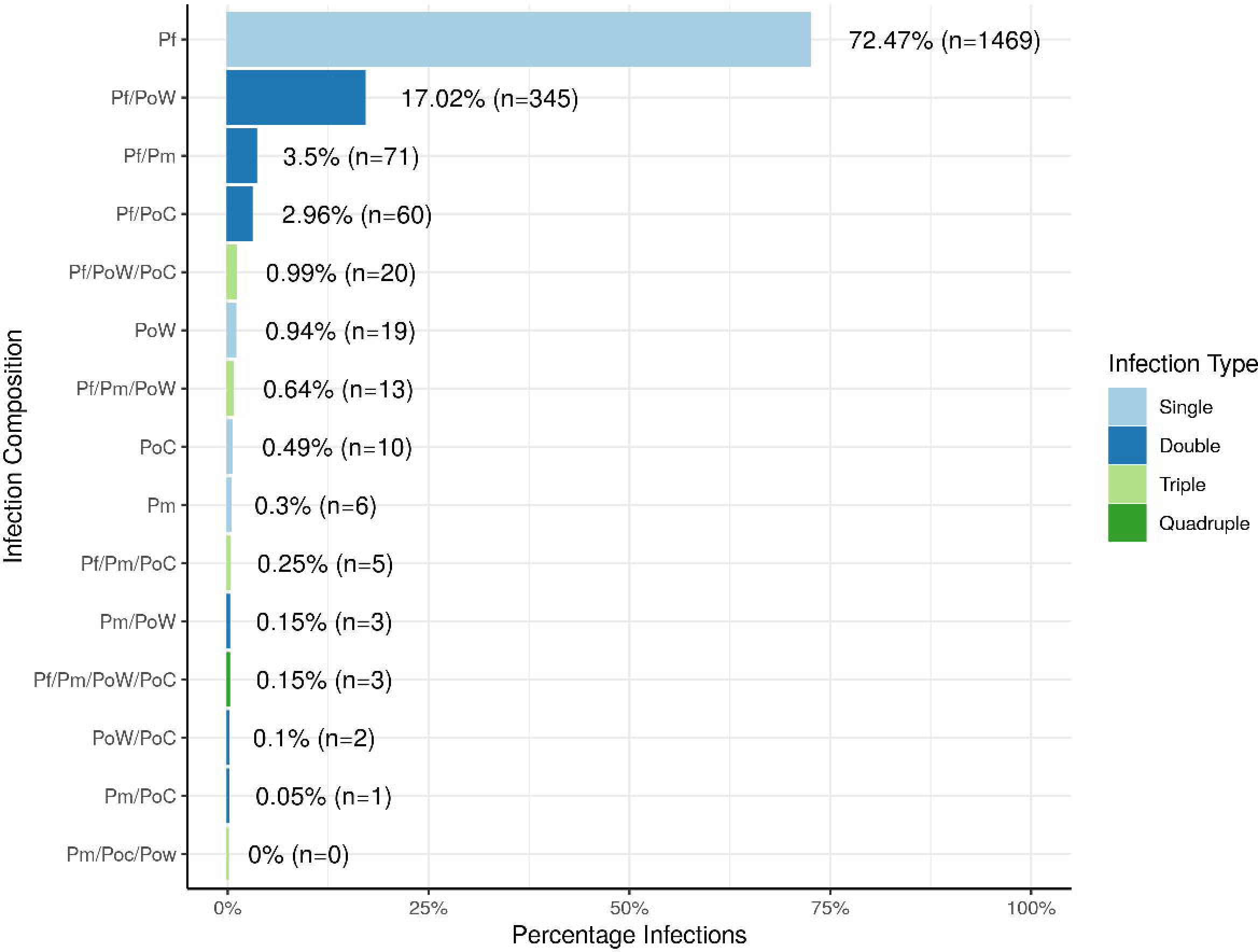
Observed Plasmodium species composition. Infections caused by only one species accounted for 74•20% of infections i.e. *P. falciparum, P. ovale wallikeri, P. ovale curtisi*, and *P. malariae*. The most prevalent multiple species infections were caused by *P. falciparum* and *P. ovale wallikeri*.

### *Plasmodium* species frequencies and inter-species interactions

Using the statistical model developed to estimate the population frequency of each *Plasmodium* species under the assumption of independent strain acquisition, i.e. no inter-species interactions, a population frequency of 89•6%, 7•1%, 1•7%, and 1•6% for *P. falciparum, P. ovale wallikeri, P. malariae* and *P. ovale curtisi*, respectively was predicted, with a mean number of extant infections equal to 2•94 (Table 2). These estimates accurately captured the observed frequency distribution of the composition of infecting species, with the 95% bootstrapped quantiles generated from these estimates containing the observed data for each infection type (Figure 3). The accuracy of the model predictions varied between infection type, with occurrence of *P. ovale wallikeri* single species infections, and *P. falciparum/P. ovale curtisi* infections being over estimated. Conversely, the model under-predicted the occurrence of *P. ovale curtisi* single species infections and *P falciparum/P. ovale wallikeri* infections. The variation in the observed distribution of infection types was better explained using a model that included interactions between *Plasmodium* species (Table 2, Figure 4), with the best fitting interaction model including one interaction term 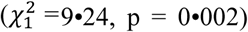, which predicted a significant interference between *P. falciparum* and *P. ovale curtisi* (k_1,3_ = 0•405). In all equivalent models, the use of a Poisson distribution to describe the number of infections yielded more parsimonious models (Supplementary Table 2), with the gradient in the model likelihood being largely flat with respect to the dispersion parameter *r* (Supplementary Figure 2). The model parameters and likelihoods of the best fitting models are shown in Supplementary Table 2.

**Table 2:** Description of each best fitting model and parameter estimates.

| Model Description |  | Model Performance |  |  | Shared Parameters |  |  |  |  | Additional Parameters |  |  |  |  |  |  |
| --- | --- | --- | --- | --- | --- | --- | --- | --- | --- | --- | --- | --- | --- | --- | --- | --- |
| Name | $Pr(\mu\vert \theta)$ | AICc | ΔAIC | LogLik | pf | pm | poc | pow | $\mu$ | $r$ | $k_{12}$ | $k_{13}$ | $k_{14}$ | $k_{23}$ | $k_{24}$ | $k_{34}$ |
| $k_{13}$<br>Interference | Poisson | 74•113 | 0 | -31•036 | 0•892 | 0•016 | 0•025 | 0•067 | 3•062 | | | 0•405 | | | | |
| $k_{13}$<br>Interference | Negative<br>Binomial | 76•190 | 2•064 | -31•067 | 0•893 | 0•016 | 0•025 | 0•067 | 3•074 | 100•001 | | 0•405 | | | | |
| Independent | Poisson | 81•346 | 7•245 | -35•658 | 0•895 | 0•017 | 0•017 | 0•072 | 2•927 |  |  |  |  |  |  |  |
| Complete<br>Interference | Poisson | 82•067 | 7•865 | -29•968 | 0•896 | 0•017 | 0•016 | 0•071 | **2•938** |  | 0•901 | 0•482 | 1•982 | 1•189 | 1•965 | 1•736 |
| Independent | Negative<br>Binomial | 83•293 | 9•180 | -35•626 | 0•909 | 0•019 | 0•026 | 0•047 | 2•759 | 99•996 |  |  |  |  |  |  |
| Complete<br>Interference | Negative<br>Binomial | 84•008 | 9•782 | -29•927 | 0•909 | 0•019 | 0•025 | 0•047 | 2•775 | 99•997 | 0•833 | 0•487 | 1•949 | 1•27 | 1•951 | 1•71 |

Each model shares parameters for the frequency of *P. falciparum* (pf), *P. malariae* (pm), *P. ovale curtisi* (poc), *P. ovale wallikeri* (pow), and the mean number of extant infections ( $\mu$ ).

**Figure 3:**
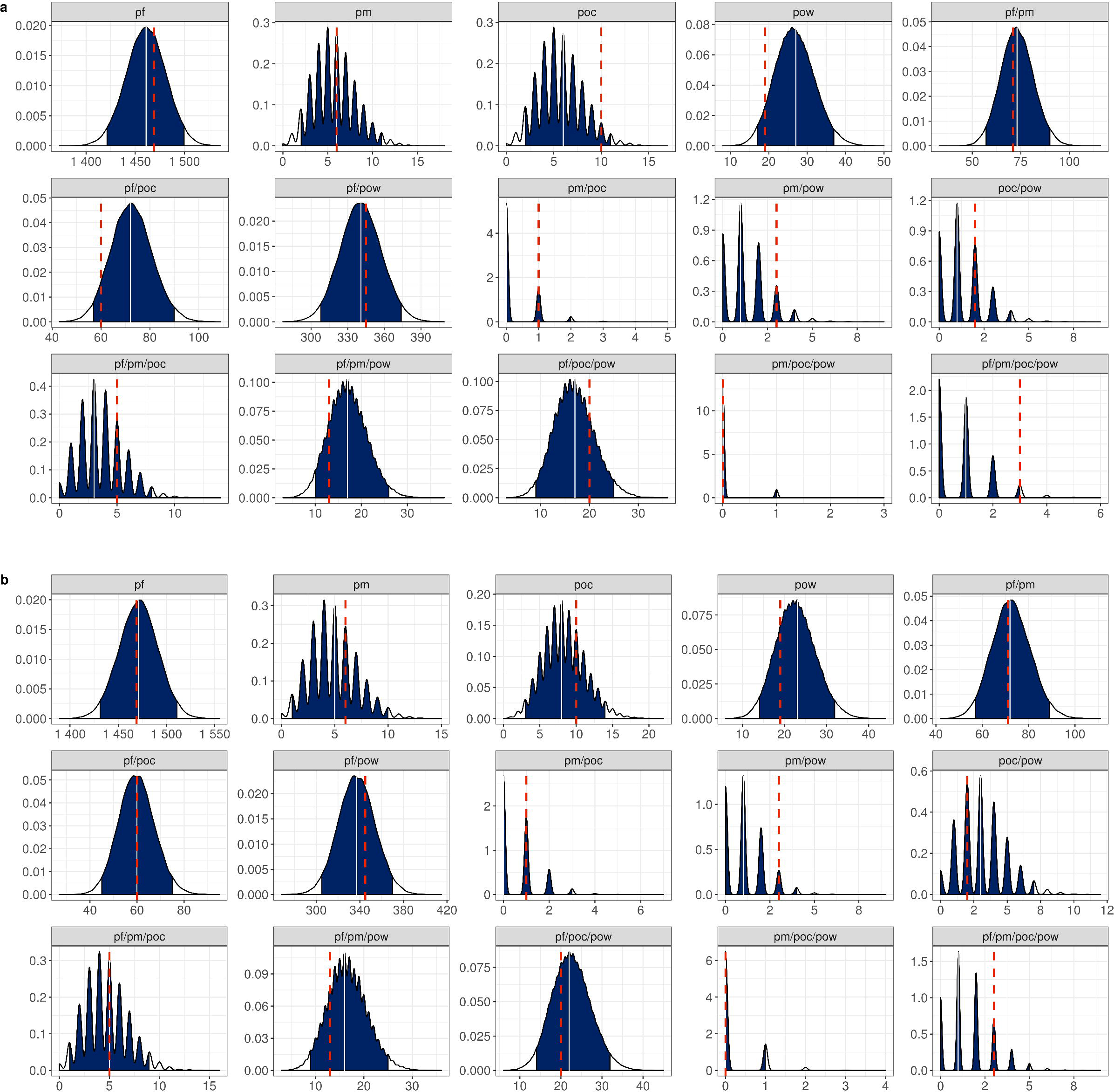
Predicted infection species composition. Plots show the estimated distribution for each infection composition, consisting of *P. falciparum* (pf), *P. malariae* (pm), *P. ovale curtisi* (poc), and *P. ovale wallikeri* (pow). Distributions were estimated using 50,000 sampling repetitions drawn from the best fitting **a)** independent and **b)** interference model. Blue regions show the 95% quantile interval, with the median shown in white line. The observed infection composition from the data is shown with the red dashed line. The interference model shown in **b)** included one additional parameter, which was an interference between *P. falciparum* and *P. ovale curtisi*.

**Figure 4:**
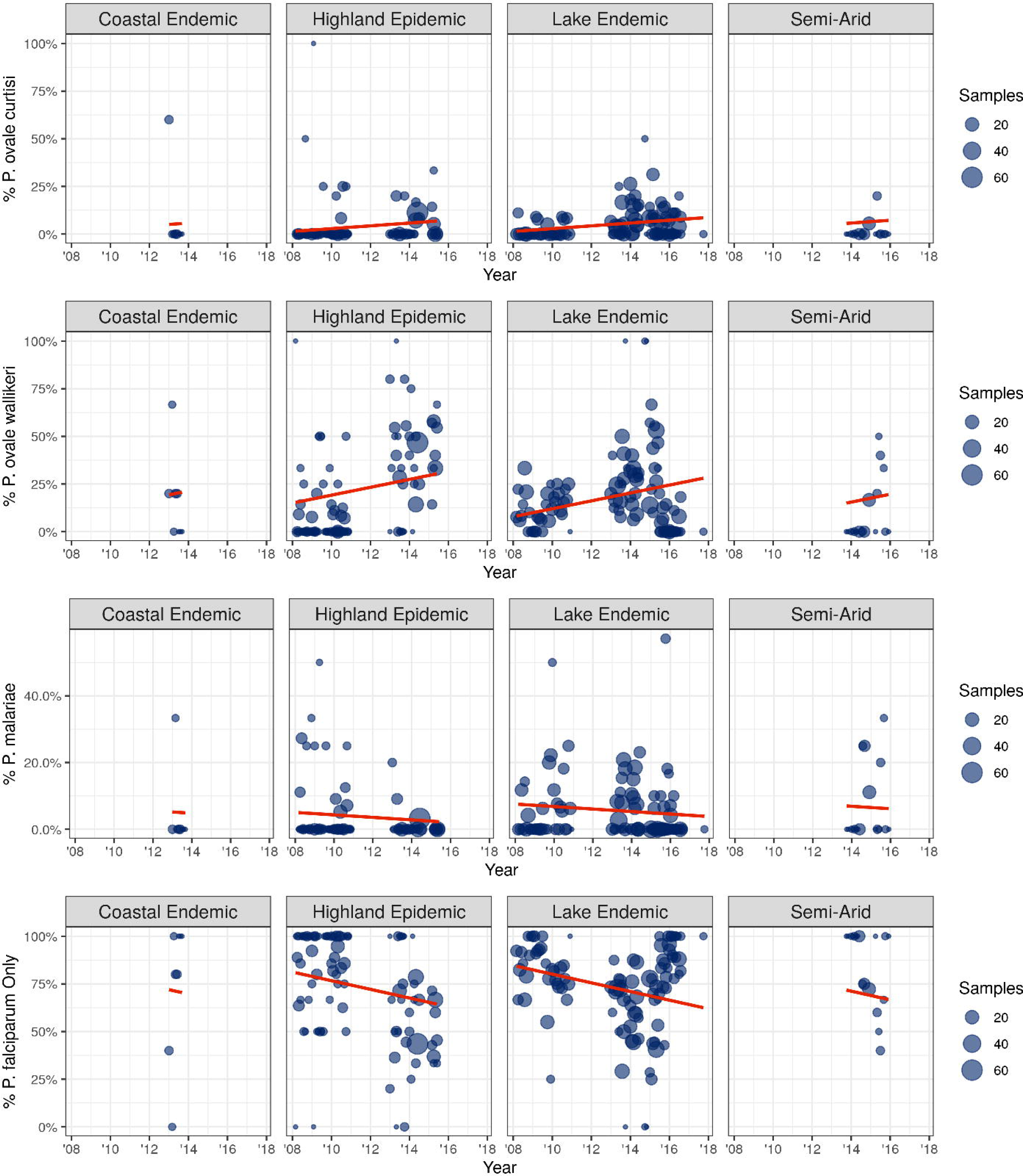
Frequency of infections containing *P. ovale curtisi, P. ovale wallikeri, P. malariae*, and only *P*. *falciparum*. Each plot shows the percentage of infections that were positive for each *Plasmodium spp*. over time for the four transmission zones sampled. A mixed-effects linear regression model with a random intercept for each transmission region was fitted to the data and is plotted in red in each plot.

### Species frequency over time

Analyses of the longitudinal trends of the infecting species composition showed significant changes occurred over the study period (Table 3). The mixed-effect linear modelling identified a significant increase in *P. ovale* spp. infections over time (*p* < 0•001), with *P. ovale wallikeri* having the largest increase (Figure 4). Conversely, there was a decrease in the frequency of infections containing *P. malariae* (*p =* 0•079) and single species infections caused by *P. falciparum* (*p* < 0•001). These findings are summarized in Table 3 and Figure 4.

**Table 3:**
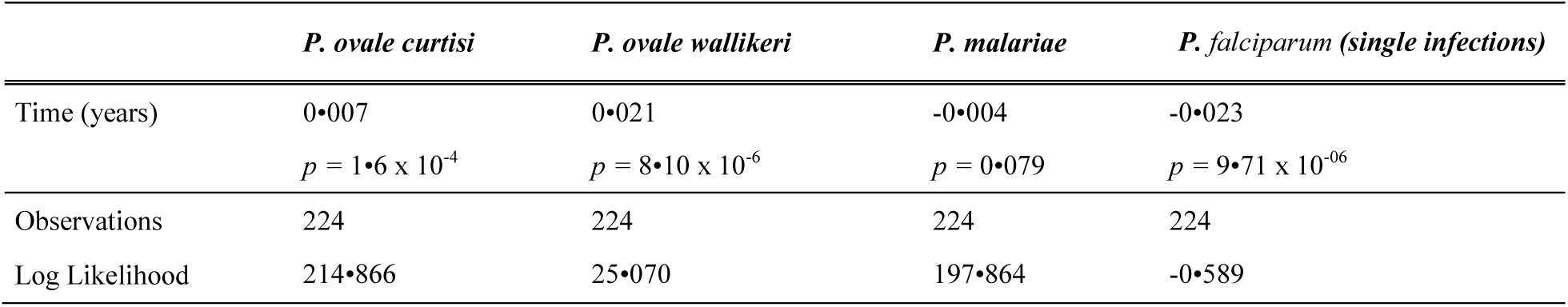
Parameters for the mixed effect analysis of *Plasmodium spp*. frequency changes over time

### Travel and infection history

Travel history indicated that Kericho (highland epidemic) had the highest number of malaria cases acquired outside the region, with 84•0% of study participants admitting that they had travelled in the last 2-4 weeks prior to becoming ill, and 93•2% of those who traveled having visited western Kenya. In contrast, only 16•3% of participants from Kisii (which is also in highland epidemic) reported having traveled, with 94•7% of those who traveled having visited western Kenya. Only a small percentage of participants in Malindi (coastal endemic) and Marigat (semi-arid) reported having traveled to western Kenya in the last 2-4 weeks prior to becoming ill. The majority of the participants in Kombewa, Kisii, and Kisumu (95•6%, 89•5%, and 86•7%, respectively) reported that they had previously contracted malaria compared to less than 60•0% of participants in Malindi, Marigat, and Kericho. This data suggests that clinical malaria burden in Kericho is majorly not due to local malaria transmission. Indeed, 72•0% (n=215) of the study participants in Kericho indicated their home of origin is western Kenya, indicating they immigrated to Kericho due to work or business, and they frequently visit their home of origin as indicated by their travel history. Further, of these, 64•7%, (n=139) reported having lived in Kericho for less than 5 years. The clinicians in Kericho noted most of the locals report to the hospital with severe malaria and thus do not qualify to be enrolled in our study. Travel and infection history data is summarized in Table 4.

**Table 4:** Study site demographic and clinical summary

| Site | Age | Sex (% female) | Fever | Malaria episodes in last 12 months | Previous Malaria Infection | Travel in last 2-4 weeks | N |
| --- | --- | --- | --- | --- | --- | --- | --- |
| Kericho | 12.25 [10.33-14.56] | 46.40% [37.60-55.20] | 73.55% [64.46-80.17] | 0.73 [ 0.56- 0.92] | 53.60% [44.00-61.60] | 84.00% [76.00-88.80] | 170 |
| Kisii | 8.49 [ 6.96-10.47] | 52.29% [44.44-59.48] | 88.89% [82.35-92.81] | 0.50 [ 0.42- 0.59] | 89.54% [83.01-92.81] | 16.34% [10.46-22.22] | 432 |
| Kisumu | 8.85 [ 7.99- 9.70] | 50.37% [45.76-54.58] | 62.31% [57.84-65.67] | 0.95 [ 0.85- 1.05] | 86.72% [83.21-89.30] | 45.20% [40.95-49.45] | 887 |
| Kombewa | 8.16 [ 7.12- 9.50] | 52.19% [46.03-56.90] | 66.44% [60.27-71.23] | 1.98 [ 1.78- 2.15] | 95.62% [92.26-97.31] | 11.45% [ 8.08-15.15] | 437 |
| Malindi | 14.36 [ 9.18-22.55] | 50.00% [22.73-68.18] | 50.00% [27.27-68.18] | 0.08 [ 0.00- 0.21] | 9.09% [ 0.00-18.18] | 50.00% [27.27-63.64] | 24 |
| Marigat | 17.71 [13.30-23.81] | 57.14% [28.57-71.43] | 71.43% [47.62-85.71] | 0.10 [ 0.04- 0.19] | 57.14% [28.57-71.43] | 33.33% [14.29-52.38] | 77 |
| All | 9.27 [ 8.75- 9.76] | 50.78% [48.28-53.10] | 68.03% [65.68-70.22] | 1.02 [ 0.96- 1.08] | 83.79% [81.98-85.69] | 36.81% [34.66-39.05] | 2027 |

\* Mean [95% Bootstrapped Confidence Interval]

### Fever presentation and symptomatic complaints

Overall, 68•0% of infections presented with fever (recorded with a temperature above 37•5°C) (Table 4). The proportion of cases presenting with fever at clinics was highest in younger individuals and decreased with increasing age (Figure 5). Kisii had the highest number of participants presenting with fever (88•9%) and Malindi had the lowest proportion of fevers (50•0%). Analysis of the risk factors associated with fever presentation in individuals positive for *P. falciparum* revealed that the infection composition was associated with fever. The odds of presenting with fever at the clinic (adjusted for age, sex, year, and previous malaria attacks) was estimated to be 0•43 times less likely if the individual was co-infected with *P. malariae* (adjusted OR: 0•43, 95% CI: 0•25 −0•74, *p = 0•0023*) (Figure 6). Additionally, fever increased over time (adjusted OR for an increase of 1 year: 1•08, 95% CI: 1•02 - 1•15), and was less prevalent in older individuals (adjusted OR for an increase of 1 year in age: 0•95, 95% CI 0•94 - 0•96).

**Figure 5:**
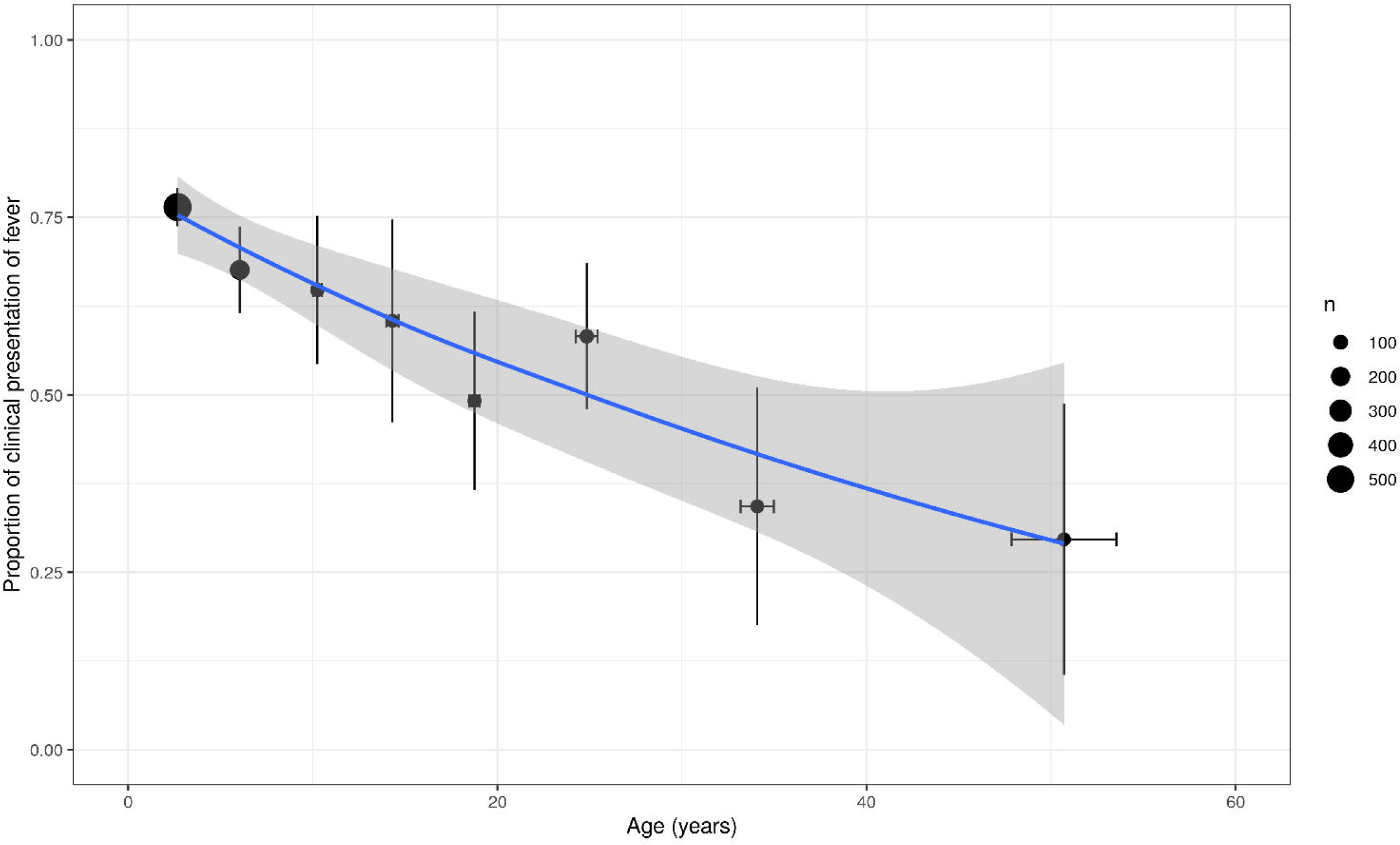
Relationship between fever presentation and age. The proportion of cases presenting with fever at clinic decreases with age. Each point represents the mean age and proportion binned within age groups and the size of point shows the number of individuals in the age binned class. The vertical and horizontal whiskers show the 95% confidence interval and a smoothed locally weighted regression with confidence intervals is shown in blue and grey.

**Figure 6:**
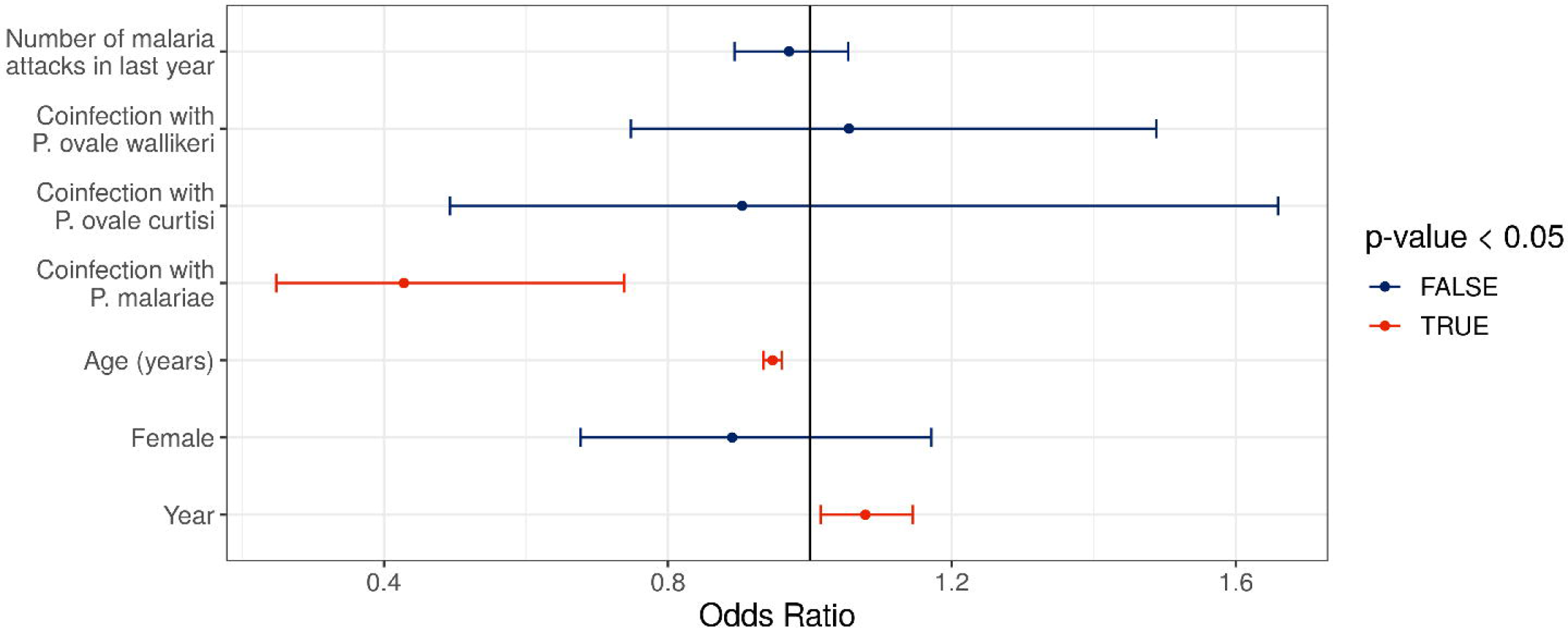
Risk factors associated with clinical presentation of fever. The odds ratio for each predictor assessed is shown with their 95% confidence intervals as whiskers surrounding each point. Odds ratios significantly not equal to 1 are shown in red and were observed for the age, year of sample collection and coinfection with *P. malariae*.

We investigated the relationship between the reported chief complaint symptoms and the malaria species infection composition of infected individuals upon arrival at clinics. Infections induced by single or mixed non-falciparum parasites induced similar symptoms as those seen with *P. falciparum* as single or mixed infections. There was one case where a *P. ovale wallikeri* single infection induced seizure in a 6-year old boy, which was also associated with severe malaria. Figure 7 shows the details on the infecting species composition and the symptoms manifested in the study participants. Lastly, we assessed whether the observed infecting species composition for each symptomatic complaint was predicted by the frequency of each malaria species estimated earlier in Table 2. Overall, the observed infection compositions occurred as expected for each symptom except for fever, for which *P. falciparum* single infections and *P. falciparum/P. ovale wallikeri* double infections did not occur within the 95% prediction quantile (see Supplementary Figure 3).

**Figure 7:**
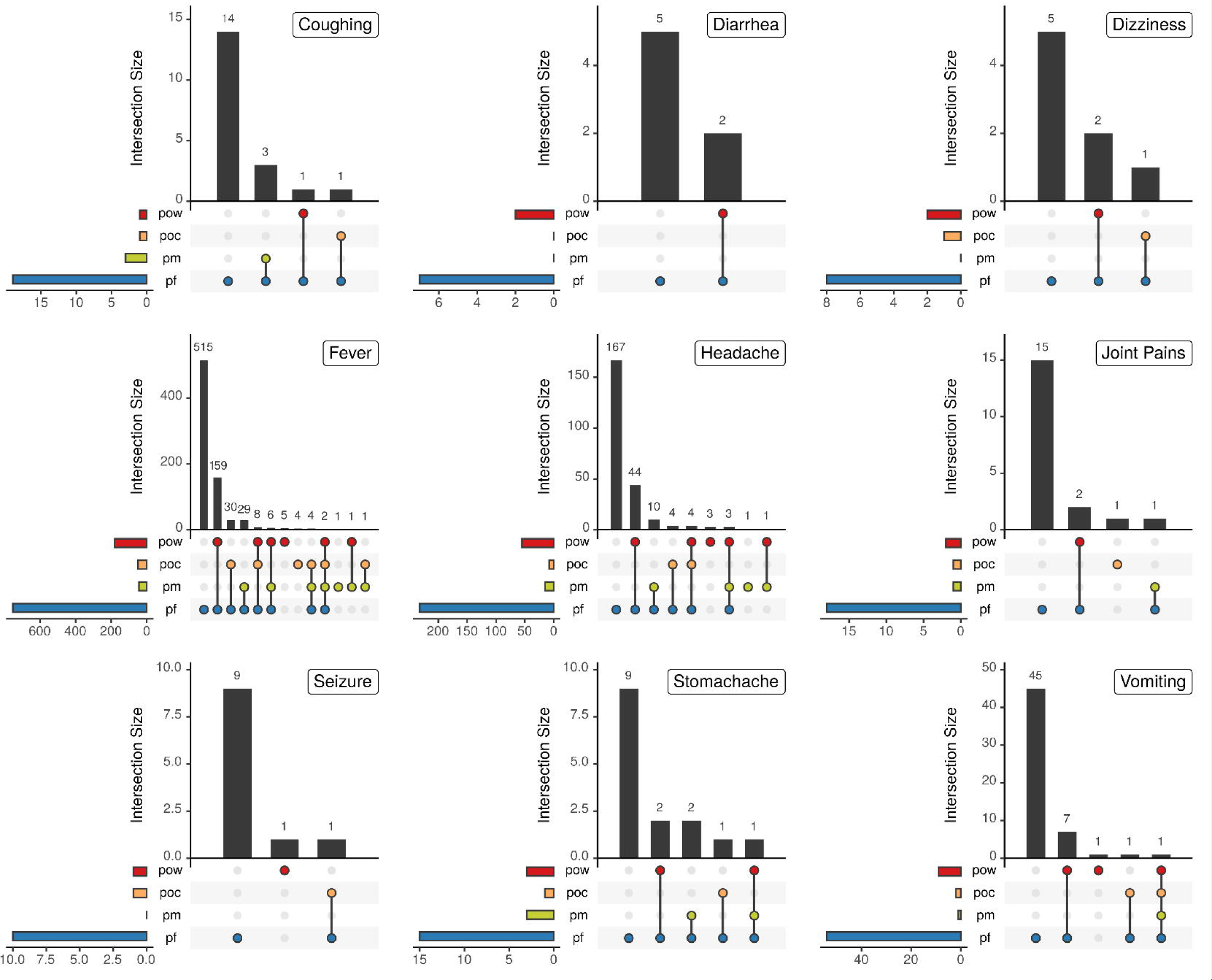
Chief complaint on arrival at clinic. The chief complaint of infected individuals upon arrival at clinic is shown with respect to the infecting species composition, consisting of *P. falciparum* (pf), *P. malariae* (pm), *P. ovale curtisi* (poc), and *P. ovale wallikeri* (pow), for all complaints reported in more than five individuals. One individual (pf) reported jaundice, one (pf) reported nausea, and five (3 pf and 2 pf/pow) reported backache.

## Discussion

Numerous studies have explored how *P. falciparum* populations are being disrupted by improved malaria control and elimination efforts. However, there are a limited number of studies that have investigated the burden, epidemiology and clinical implication of *P. ovale* spp. and *P. malariae* in malaria endemic settings. In this study we conducted a longitudinal field study in six sites in Kenya, recording the clinical presentation and infection composition of clinical malaria episodes, which allowed the prevalence of non-falciparum malaria to be estimated. This study was conducted over a period of eight years, which spans the introduction of ACT in Kenya as the first-line treatment for uncomplicated malaria. Using novel statistical models to estimate the frequency of each malaria species, we estimate a higher frequency of non-falciparum species than previously shown. Over the study period, there was a significant increase in infections containing non-falciparum malaria, and a significant decrease in infections containing *P. falciparum* single infections. These observations suggest that the decrease in malaria prevalence exhibited over the study period has changed the composition of *Plasmodium* spp. circulating in these settings. The implications of this disruption in the context of treatment, control and elimination effort must be addressed.

There was a wide array of malaria symptoms recorded as the chief complaints when the study participants were interviewed at the clinics. Majorly, fever and headache were the chief complaints followed by vomiting, and coughing. Joint pains and backaches were also reported but mostly in older study participants. Clinical episodes due to non-falciparum malaria infections caused similar symptoms as those caused by *P. falciparum* malaria, with seizure reported by some, including those infected only with non-falciparum malaria. Overall, the proportion of patients with fever decreased with age. A larger proportion of patients from the highland epidemic zone had fever at the time of enrollment compared to those from the endemic zones, especially children <5 years old which may indicate lower population immune status. Additionally, *P. falciparum* coinfection with *P. malariae* was significantly associated with a decreased risk of presenting with fever at clinic. This is in agreement with previous studies in Nigeria and Ghana that have also shown decreased overt malaria clinical presentation in coinfections of *P. falciparum* and *P. malariae.^18, 21^*

*Our findings showed that during the study period, there was a significant increase in the proportion of infections carrying P. ovale wallikeri* and *P. ovale curtisi*, but a decrease in infections carrying *P. malariae* and *P. falciparum* as single species. *P. ovale wallikeri* was the most prevalent non-falciparum species across all regions and showed the largest proportional increase over the study period. The statistical modelling developed confirmed this, estimating the population frequency of *P. ovale wallikeri* to be 6•7% compared to 2•5% for *P. ovale curtisi*. These estimates were produced using the overall best fitting model, which also suggested a significant interference between *P. falciparum* and *P. ovale curtisi*, which approximately halves the probability of a successful subsequent infection by *P. ovale curtisi* after an initial infection by *P. falciparum* and vice versa. This interaction could also explain the greater increase in *P. ovale wallikeri* over time compared to *P. ovale curtisi* and lends support to hypotheses that *P. ovale wallikeri* and *P. ovale curtisi* have different within-host adaptations.^2^

An alternative explanation for the increase in *P. ovale* spp. could be due to the differential impact of AL on *P. ovale* spp. This may lead to unresolved infections following AL treatment, which have been previously reported.^7, 10, 11^ In a longitudinal study conducted in Tanzania, *P. falciparum* declined over the study period but the prevalence of *P. ovale* spp. and *P. malariae* increased 6- and 2-fold, respectively.^22^ This study also coincided with the introduction of AL in Tanzania. In our study, we observed an increase in the frequency of *P. ovale* spp. (but not *P. malariae*), coinciding with the introduction of AL in Kenya. This might be due to several factors including unresolved infections following AL treatment^10, 11, 23^ and/or relapsing malaria.^23-25^ It is also possible that *P. ovale* spp. response to AL is becoming attenuated over time. Additional studies are required to further investigate how the use of AL and other ACTs might be playing a role in the shift of *Plasmodium* spp. composition in Kenya and elsewhere in sSA.

There are a number of limitations in our study. Firstly, the samples analyzed by this study were obtained from symptomatic individuals which may not be representative of the malaria prevalence in the population as a whole. In addition, in the statistical modelling for estimating the frequencies of *Plasmodium* species, we assumed that each acquired strain was sequentially acquired due to successive bites. However, it is possible that multiple species could be passed on within one infectious bite. This could be an alternative explanation for the differences seen between the observed and predicted infection types using the independent model (Figure 3). For example, this could not be due to between species interactions within the host, but due to an increased affinity or differing adaptation to the vector, with *P. falciparum*, and *P. ovale wallikeri* able to be passed on within the same infectious bite. Despite this, predictions from the best fitting model were highly accurate and the developed methodology is flexible enough to enable alternative models for the likelihood of any given infection type. Lastly, although samples were collected from six sites spanning four transmission zones, the longitudinal coverage of samples from the coastal endemic and semi-arid zone was substantially less than the other regions. Conclusions drawn relating to the rate of change of *Plasmodium* species over time are thus largely informed by the highland epidemic and lake epidemic zones. The altered endemicity of these zones could subsequently be driving the patterns seen, with the increased seasonality in these epidemic regions altering the presentation of mixed-species infections.

In conclusion, the frequency of non-falciparum species collected between 2008 and 2016 was comparable across all the four transmission zones in Kenya. Our developed statistical model for estimating the frequency of *Plasmodium* species and between species interactions predicted a significant interference between *P. falciparum* and *P. ovale curtisi*, which would be, to our knowledge, one of the first efforts to statistically show between species interactions. Additionally, the risk of *P. falciparum* infections presenting with fever was 0•43 time less likely if co-infected with *P. malariae*. Lastly, the proportion of infections that were positive for infection by *P. ovale wallikeri* and *P. ovale curtisi* was observed to significantly increase over the period of study and could be as a result of the increased effective drug pressure exerted on *Plasmodium* species that do not possess a dormant hypnozoite stage. The observed increase in dormant *Plasmodium* infections could thus explain the increased observation of traveler malaria originating from Kenya and other malaria endemic areas that use ACTs. Increased surveillance for non-falciparum species infections is recommended within both symptomatic and asymptomatic individuals to monitor the changing risk of malaria infection from non-falciparum species.

## Data Availability

All data generated by this study was was available to the authors. The data is also available for this submission

## Acknowledgments

We thank Dr Veronica Manduku, KEMRI Center for Clinical Research; LTC Claire A Cornelius, Dr Douglas Shaffer, Directors, USAMRD-A/K, and Dr. Steve Munga, KEMRI Center for Global Health Research, for supporting this study and giving their permission to publish these data. We also thank all clinical staff at Kisumu East District Hospitals for their assistance.

## Disclaimer

Material has been reviewed by the Walter Reed Army Institute of Research. There is no objection to its presentation and/or publication. The opinions or assertions contained herein are the private views of the author, and are not to be construed as official, or as reflecting true views of the Department of the Army or the Department of Defense. The investigators have adhered to the policies for protection of human subjects as prescribed in AR 70–25.

Financial support: Funding for this study was provided by the Armed Forces Health Surveillance Branch (AFHSB) and its Global Emerging Infections Surveillance (GEIS) Section, Grant P0209_15_KY. The study sponsor had no role in study design; in the collection, analysis, and interpretation of data; in the writing of the report; and in the decision to submit the paper for publication. The corresponding author should confirm that he or she had full access to all the data in the study and had final responsibility for the decision to submit for publication.

## Supplementary Material

### Supplementary Methods

#### Estimating the frequency of *Plasmodium* species and assessing inter-species interaction

Let *S* = {*s*_1_, *s*_2_, *s*_3_, *s*_4_}*f* be the set of all species of interest, where *s*_1_ corresponds to *P. falciparum, s*_2_ to *P. malariae, s*_3_ to *P. ovale curtisi*, and *s*_4_ to *P. ovale wallikeri*. We assume that each successful infectious bite from a mosquito passes on one of these parasite species to a human host, which is draw.00……..n at random from *S* with probabilities *P* = {*p*_1_, *p*_2_, *p*_3_, *p*_4_} respectively, where 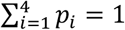. The unobserved sequence of species passed to a host can be written *Y* = {*y*_1_, *y*_2_, …, *y*_*k*_} ∈ *S*, where *μ* is the total number of infections this host receives. We can also write this in frequency form as *N*_*Y*_ = {*n*_1_, *n*_2_, *n*_3_, *n*_4_}, where 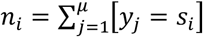. The probability of the unobserved data is therefore multinomial, and can be written:

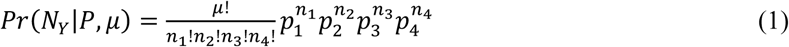

We assume that multiple inoculations of the same species are indistinguishable in the observed data – in other words the observed data consists of the set of unique species in *Y*. If *X* denotes the observed data then we can define, *X* = {*s*_*i*_: *s*_*i*_ ∈ *Y*}, |*X*| ≤ *μ*. The probability of the observed data can be obtained by summing over the individual probabilities of all unobserved sequences that are consistent with *X*, that is, they consist of *µ* infections and contain *X* as a subset. We can write:

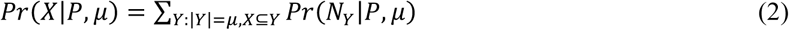

We can remove the dependency on *μ* by assuming a particular distribution for the number of infections per host. We will explore several models for this distribution below, but in each case, we can marginalise over *µ* as follows:

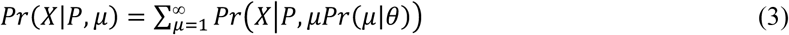

Where *θ* is a vector of parameters governing the shape of the distribution over *μ*. Finally, we note that the total number of possible values of *X*, is equal to 2^|*S*|^ - 1, which represents the sum of all non*-*empty subsets of *S*. This is equivalent to the power set of *S, P*(*S*), without the empty set, which leads to just 15 possible values for the four-species case here. Hence, the observed data can be written efficiently in the form of a table of the number of times each combination of infections was observed (see Table 3 for the equivalent table). If the values in this table are denoted 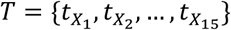 for each of the possible sets *X*_1_ to *X*_15_, then the overall probability of the data (the likelihood) is multinomial with probabilities taken from (3), and can be written as follows:

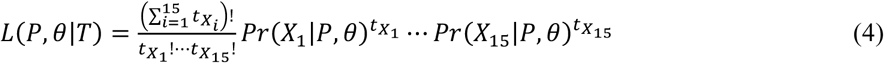

We used the likelihood in (4) to generate maximum likelihood parameter estimates for 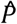 and 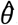. The likelihood was maximized using box-constrained optimisation ^27^ in R ^28^. The parameters 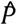 and 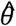 were used to simulate the model-predicted distribution for the frequency of each possible observed infection type. The estimated distribution was used to test for a difference between the expected frequencies and the observed frequencies of each infection composition using a non*-*parametric bootstrapping method. 50,000 bootstrap repetitions were drawn from the null distribution and used to estimate the median and 95% confidence interval for each infection composition, which were compared to the observed infection compositions.

We continued by extending the statistical model to test for the presence of within-host interactions between infecting *Plasmodium* species. To do this we altered our assumption that each infection is independent of previous infections and instead assume that the probability of each additional infection is dependent on the first infecting species. Let *K* = {*k*_12_, *k*_13_, *k*_14_, *k*_23_, *k*_24_, *k*_34_} be the set of between-species interactions, such that *k*_12_ is the interaction between species *s*_1_ and *s*_2_, which in our case is the interaction between *P. falciparum* and *P. malariae*. As before, we assume that the first infectious bite from a mosquito passes on one species to a human host, which is drawn at random from *S* with probabilities *P* = {*p*_1_, *p*_2_, *p*_3_, *p*_4_} respectively, where 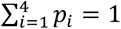. Each additional infectious bite from a mosquito passes on one species, which is now drawn at random with probabilities *Q* = {*q*_1_, *q*_2_, *q*_3_, *q*_4_}, given by:

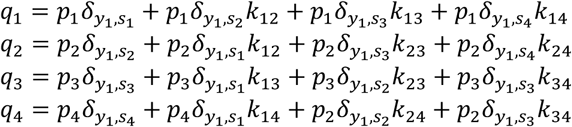

Where *y*_1_ is the first species that infected the host and 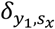 denotes the Kronecker delta (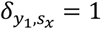 if *k*_*x,y*_ < 1, and 0 otherwise). For example, if the first species was *P. falciparum* then 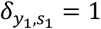 and 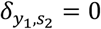. If *k*_*x,y*_ < 1, the interaction between species *x* and *y* represents a between-species interference, i.e. the presence of one species reduces the probability of the other species being acquired, which could represent inter-species competition. Conversely, If *k*_*x,y*_ > 1, the interaction term between species *x* and *y* represents between-species synergy, i.e. the presence of one species increases the probability of the other species being acquired. Lastly if *K* = {1,1,1,1,1,1}, the interference model is identical to the model of independent acquisition of strains model.

If we define *m*_*x*_ = *n*_*x*_ - *δ*_*i,x*_, the probability of the unobserved species data is given by:

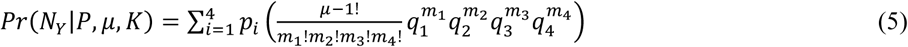

As before, we use this probability to define the probability of the observed data (equation 6) and the overall likelihood under our modelled assumption of between species interactions (equation 7) as follows:

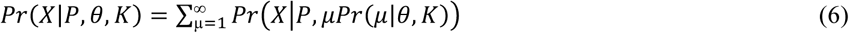

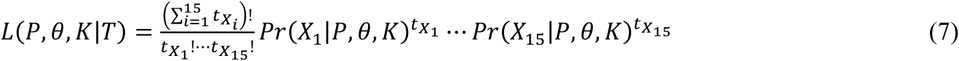

Again we maximized this likelihood and compared the predictions between the different models generated. For the model assuming species interaction, we fit one model for each of the 63 non*-*empty subsets of *K*, setting members of *K* not in a given subset set equal to 1, i.e. independent strain acquisition. For both the independent and interference model we explored both a Poisson and negative binomial distribution to describe the number of infections. The best fitting interference model was identified through comparisons of sample-size corrected Aikaike information criterion (AICc) ^18^. Finally, the best fitting interaction models were compared to the independent model using log-likelihood ratio tests to statistically test for the presence of between species interactions.

## Supplementary Results

**Supplementary Figure 1:**
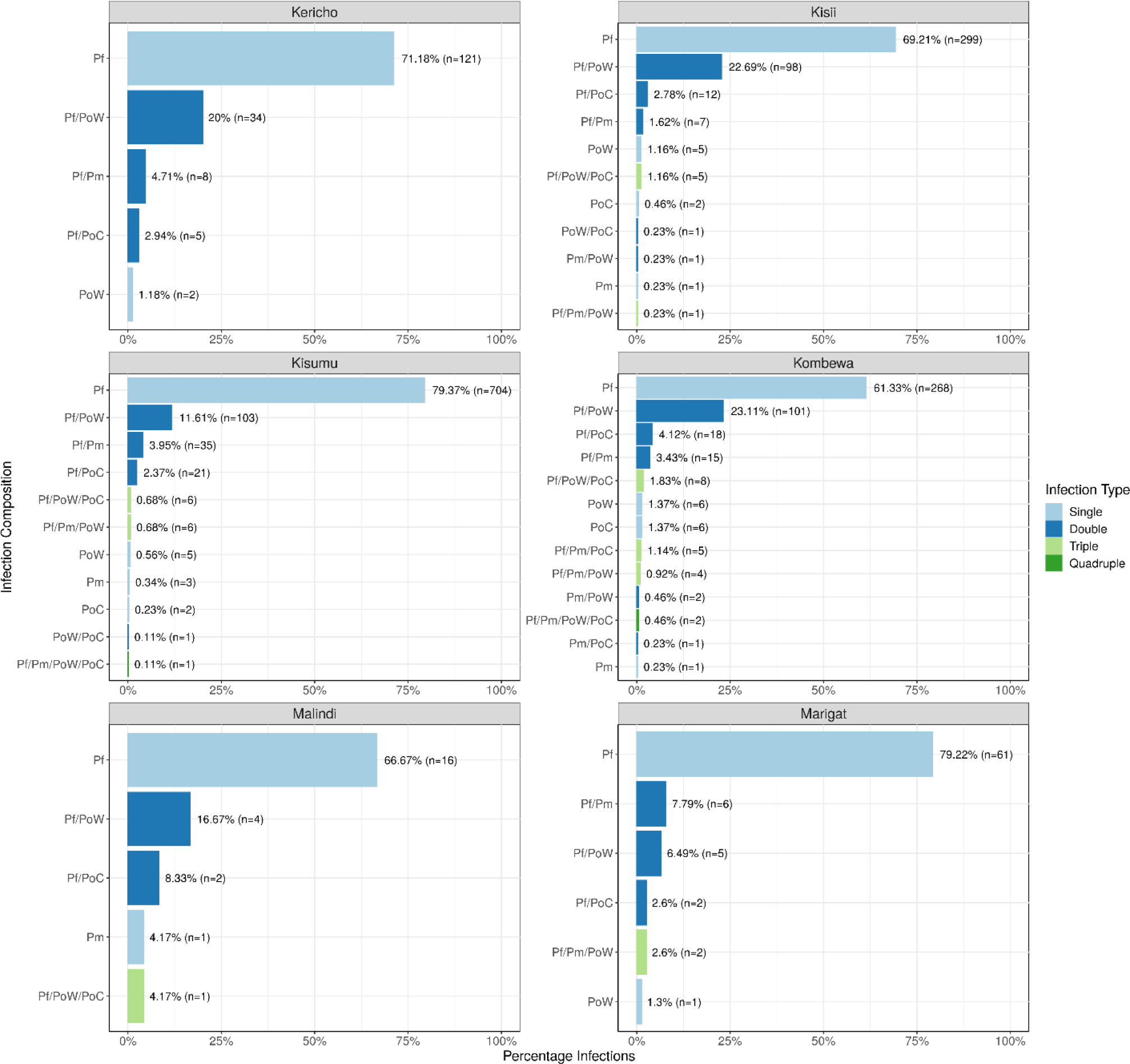
Observed Plasmodium species composition by site. Infection composition is shown for each site. *P. falciparum* single species infection was the most prevalent in each study site, followed by *P. falciparum/P. ovale wallikeri* dual infections in each site except Marigat, in which *P. falciparum/P. malariae* dual infections were the second most prevalent infection type.

**Supplementary Figure 2:**
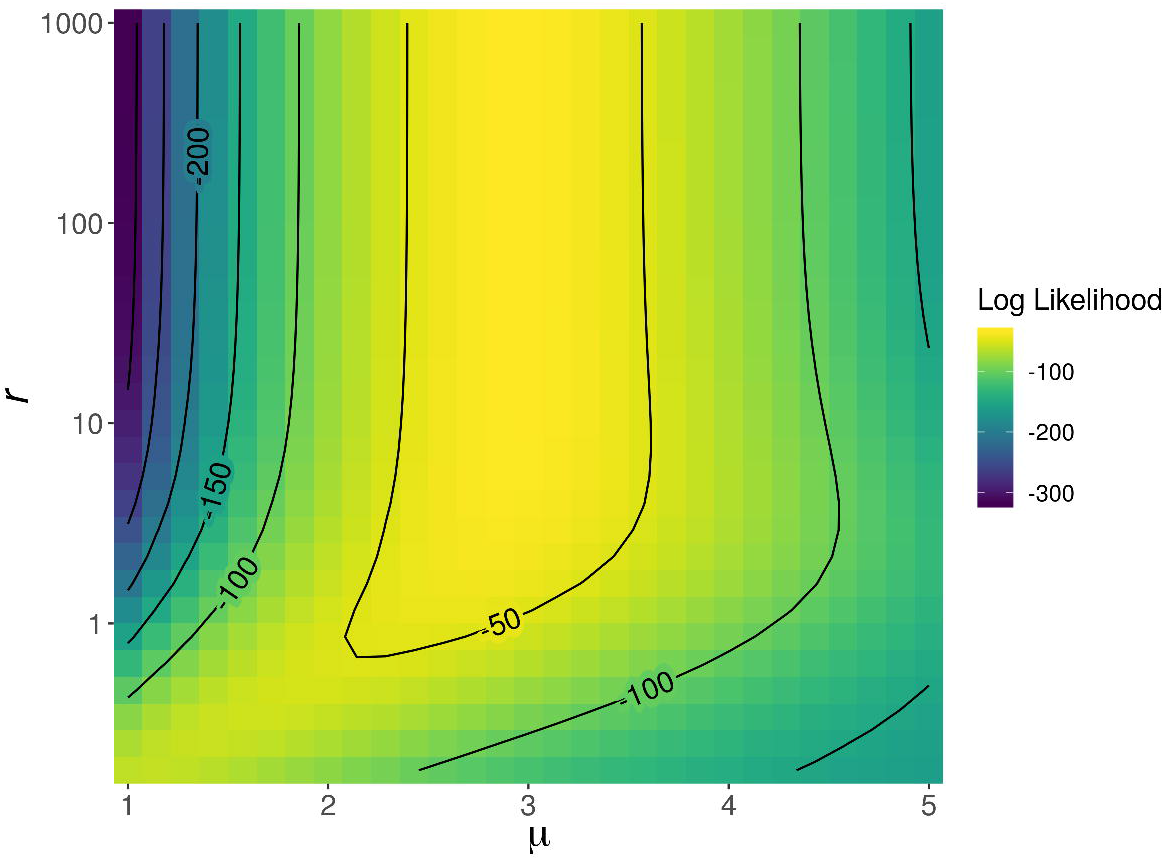
Impact of the negative binomial shape parameter on model likelihood of the interference model. The heatmap shows the relationship between the dispersion parameter, *r*, and the mean, *µ*, for the independent model with an assumed negative binomial distribution describing the number of infections. The likelihood is largely unchanged above 1 for well-chosen values of *µ*, which confirms that the distribution in the number of infections is not over-dispersed and is well explained by the Poisson distribution.

**Supplementary Figure 3:**
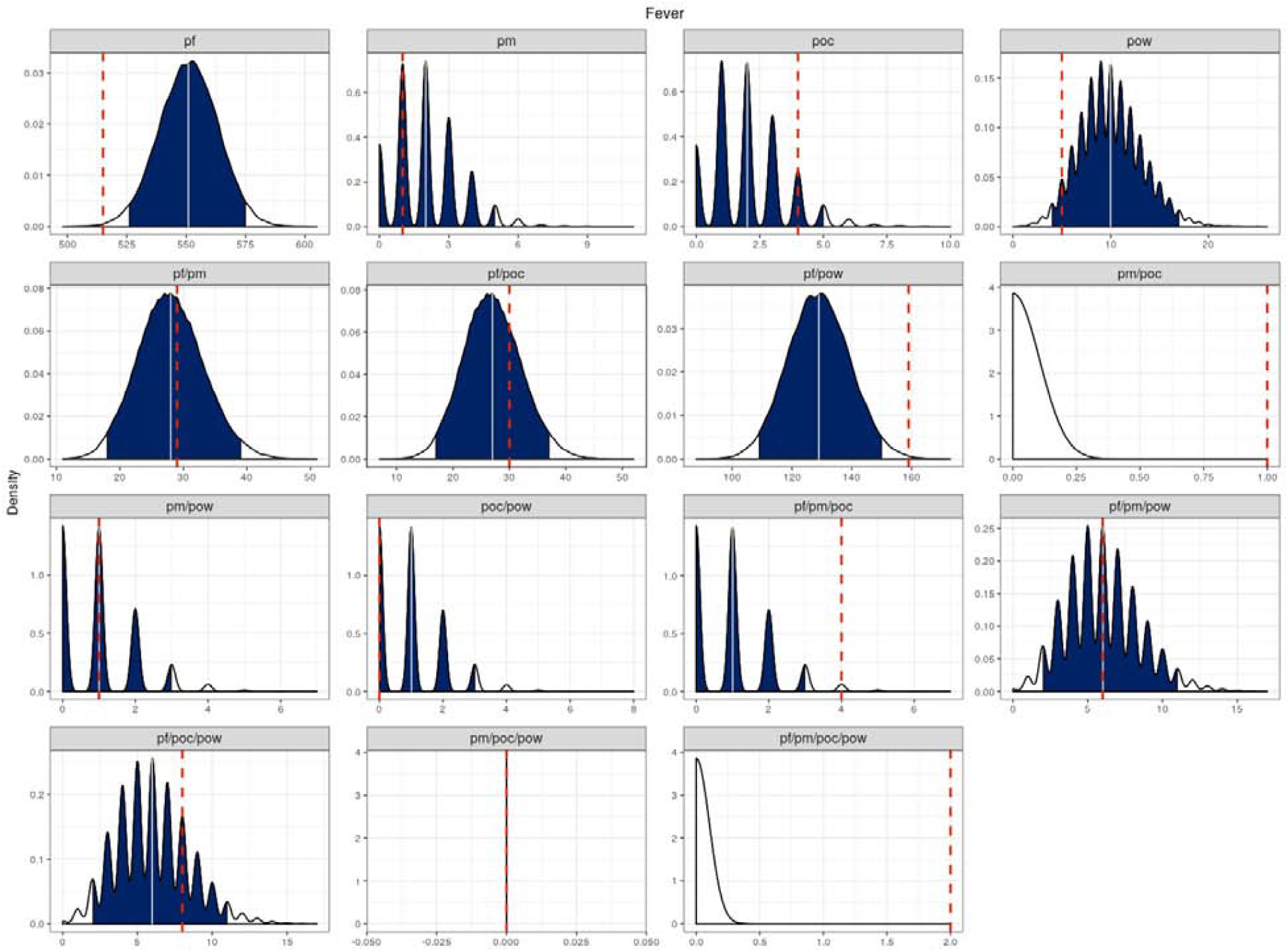
Predicted infection species composition. Plots show the estimated distribution for each infection composition, consisting of *P. falciparum* (pf), *P. malariae* (pm*), P. ovale curtisi* (poc), and *P. ovale wallikeri* (pow), for individuals whose chief complaint was reported as fever. Distributions were estimated using 50,000 sampling repetitions drawn from the best fitting model of inter-species interactions. Blue regions show the 95% quantile interval, with the median shown in white line. The observed infection composition from the data is shown with the red dashed line.

**Supplementary Table 1:**
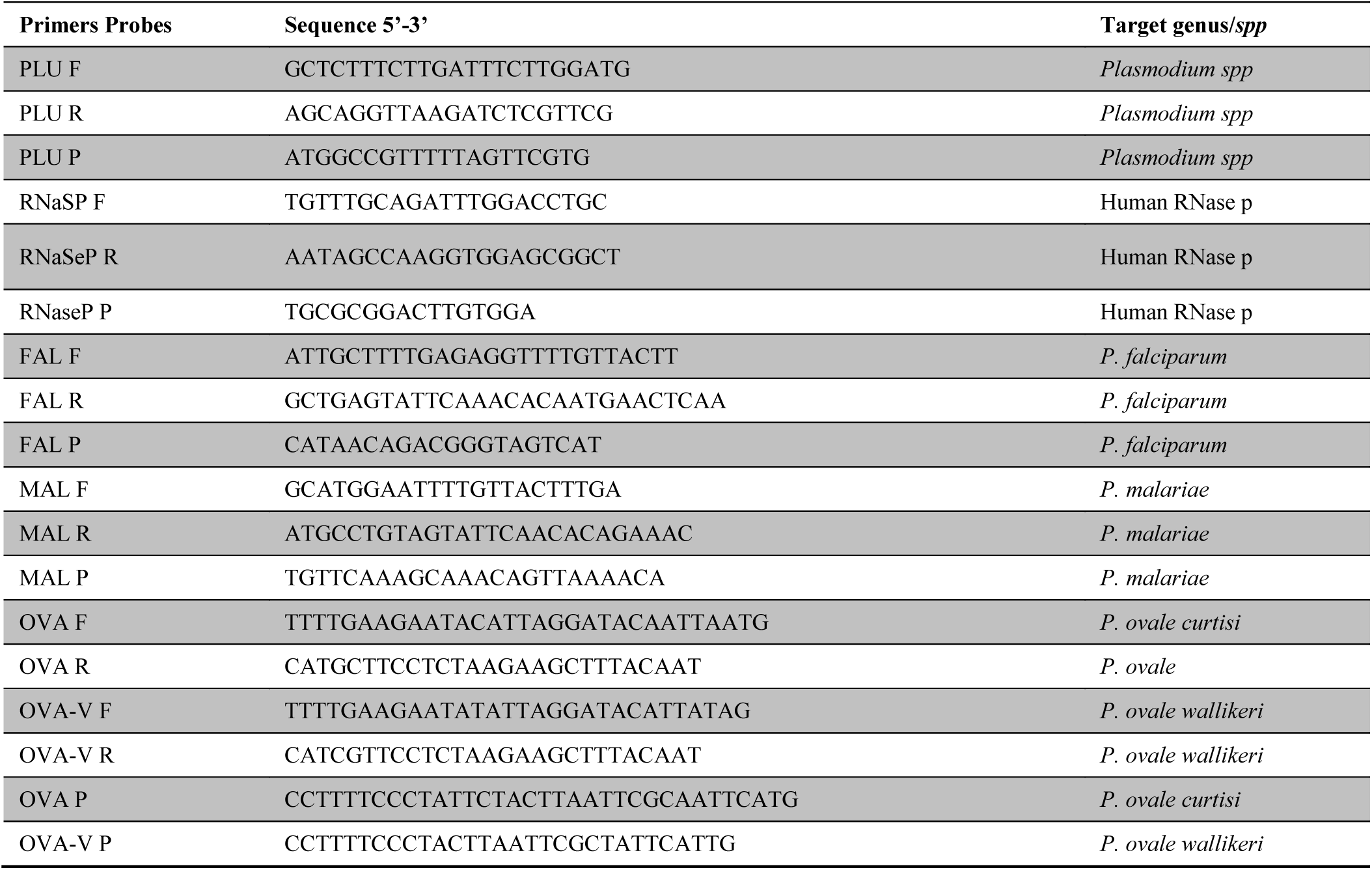
Primers and probes used for screening and identification of *Plasmodium* species ^9^.

**Supplementary Table 2:** Model Performance of all fitted models

| Model | $Pr(\mu\vert \theta)$ | AICc | AIC | $\Delta AIC$ | N. Params | LogLik |
| --- | --- | --- | --- | --- | --- | --- |
| k_13 Interference | Poisson | 74•11267 | 74•07108 | 0 | 6 | -31•0355 |
| k_12/k_13 Interference | Poisson | 74•76688 | 74•71141 | 0•640322 | 7 | -30•3557 |
| k_13/k_34 Interference | Poisson | 74•94812 | 74•89265 | 0•821568 | 7 | -30•4463 |
| k_13/k_14 Interference | Poisson | 74•99234 | 74•93686 | 0•865781 | 7 | -30•4684 |
| k_12/k_13/k_14 Interference | Poisson | 75•53879 | 75•46743 | 1•396346 | 8 | -29•7337 |
| k_13/k_23 Interference | Poisson | 75•66464 | 75•60917 | 1•538085 | 7 | -30•8046 |
| k_13/k_24 Interference | Poisson | 76•00172 | 75•94624 | 1•87516 | 7 | -30•9731 |
| k_13 Interference | Negative Binomial | 76•19009 | 76•13462 | 2•063532 | 7 | -31•0673 |
| k_12/k_13/k_34 Interference | Poisson | 76•19426 | 76•1229 | 2•051818 | 8 | -30•0615 |
| k_13/k_14/k_24 Interference | Poisson | 76•29582 | 76•22446 | 2•153377 | 8 | -30•1122 |
| k_12/k_13/k_24 Interference | Poisson | 76•43539 | 76•36404 | 2•292954 | 8 | -30•182 |
| k_12/k_13 Interference | Negative Binomial | 76•86642 | 76•79506 | 2•723977 | 8 | -30•3975 |
| k_13/k_34 Interference | Negative Binomial | 77•01882 | 76•94746 | 2•876376 | 8 | -30•4737 |
| k_13/k_14 Interference | Negative Binomial | 77•04103 | 76•96968 | 2•898593 | 8 | -30•4848 |
| k_13/k_23/k_24 Interference | Poisson | 77•32565 | 77•25429 | 3•183206 | 8 | -30•6271 |
| k_13/k_23/k_34 Interference | Poisson | 77•46203 | 77•39068 | 3•319592 | 8 | -30•6953 |
| k_12/k_13/k_14/k_23 Interference | Poisson | 77•54192 | 77•45268 | 3•381596 | 9 | -29•7263 |
| k_12/k_13/k_23 Interference | Poisson | 77•55053 | 77•47917 | 3•408086 | 8 | -30•7396 |
| k_12/k_13/k_14 Interference | Negative Binomial | 77•69272 | 77•60348 | 3•532395 | 9 | -29•8017 |
| k_13/k_23 Interference | Negative Binomial | 77•74723 | 77•67587 | 3•604791 | 8 | -30•8379 |
| k_12/k_13/k_14/k_24 Interference | Poisson | 78•10572 | 78•01648 | 3•945398 | 9 | -30•0082 |
| k_13/k_14/k_23/k_24 Interference | Poisson | 78•13243 | 78•04319 | 3•972104 | 9 | -30•0216 |
| k_12/k_13/k_14/k_34 Interference | Poisson | 78•13669 | 78•04745 | 3•976366 | 9 | -30•0237 |
| k_12/k_13/k_34 Interference | Negative Binomial | 78•27895 | 78•18971 | 4•118627 | 9 | -30•0949 |
| k_13/k_14/k_24 Interference | Negative Binomial | 78•36309 | 78•27385 | 4•202766 | 9 | -30•1369 |
| k_14/k_23/k_24/k_34 Interference | Poisson | 78•48195 | 78•39271 | 4•321625 | 9 | -30•1964 |
| k_12/k_13/k_24/k_34 Interference | Poisson | 78•57705 | 78•48781 | 4•416723 | 9 | -30•2439 |
| k_13/k_14/k_24/k_34 Interference | Poisson | 78•76873 | 78•67949 | 4•608405 | 9 | -30•3397 |
| k_12/k_13/k_23/k_34 Interference | Poisson | 78•81423 | 78•72498 | 4•6539 | 9 | -30•3625 |
| k_13/k_14/k_23 Interference | Negative Binomial | 78•87504 | 78•7858 | 4•714718 | 9 | -30•3929 |
| k_13/k_24/k_34 Interference | Negative Binomial | 78•92236 | 78•83312 | 4•76204 | 9 | -30•4166 |
| k_13/k_14/k_34 Interference | Poisson | 78•94966 | 78•8783 | 4•807219 | 8 | -31•4392 |
| k_14/k_23/k_34 Interference | Poisson | 79•32514 | 79•25378 | 5•182697 | 8 | -31•6269 |
| k_12/k_13/k_23 Interference | Negative Binomial | 79•34714 | 79•25789 | 5•18681 | 9 | -30•6289 |
| k_13/k_23/k_24/k_34 Interference | Poisson | 79•3874 | 79•29816 | 5•227078 | 9 | -30•6491 |
| k_13/k_23/k_24 Interference | Negative Binomial | 79•42411 | 79•33487 | 5•263788 | 9 | -30•6674 |
| k_12/k_13/k_14/k_23 Interference | Negative Binomial | 79•61014 | 79•50102 | 5•429932 | 10 | -29•7505 |
| k_13/k_24 Interference | Negative Binomial | 79•67027 | 79•59891 | 5•527829 | 8 | -31•7995 |
| k_13/k_14/k_23 Interference | Poisson | 79•67639 | 79•60503 | 5•533948 | 8 | -31•8025 |
| k_14/k_24/k_34 Interference | Poisson | 80•00145 | 79•93009 | 5•859004 | 8 | -31•965 |
| k_12/k_13/k_14/k_34 Interference | Negative Binomial | 80•01937 | 79•91024 | 5•839161 | 10 | -29•9551 |
| k_12/k_13/k_14/k_23/k_34 Interference | Poisson | 80•03396 | 79•92484 | 5•853752 | 10 | -29•9624 |
| k_12/k_13/k_24/k_34 Interference | Negative Binomial | 80•20901 | 80•09988 | 6•028795 | 10 | -30•0499 |
| k_13/k_14/k_23/k_24 Interference | Negative Binomial | 80•22392 | 80•11479 | 6•04371 | 10 | -30•0574 |
| k_12/k_13/k_14/k_23/k_24 Interference | Poisson | 80•25194 | 80•14281 | 6•071731 | 10 | -30•0714 |
| k_12/k_13/k_14/k_24/k_34 Interference | Poisson | 80•39552 | 80•28639 | 6•215308 | 10 | -30•1432 |
| k_13/k_14/k_23/k_24/k_34 Interference | Poisson | 80•3974 | 80•28827 | 6•217186 | 10 | -30•1441 |
| k_13/k_14/k_23/k_34 Interference | Poisson | 80•45992 | 80•37068 | 6•299592 | 9 | -31•1853 |
| k_23 Interference | Poisson | 80•56241 | 80•52083 | 6•449746 | 6 | -34•2604 |
| k_23/k_34 Interference | Poisson | 80•57998 | 80•52451 | 6•453425 | 7 | -33•2623 |
| k_12/k_14/k_34 Interference | Poisson | 80•88739 | 80•81603 | 6•744948 | 8 | -32•408 |
| k_13/k_14/k_24/k_34 Interference | Negative Binomial | 80•94324 | 80•83411 | 6•763031 | 10 | -30•4171 |
| k_13/k_14/k_34 Interference | Negative Binomial | 80•94911 | 80•85987 | 6•788784 | 9 | -31•4299 |
| k_12/k_13/k_23/k_24 Interference | Poisson | 80•98562 | 80•89638 | 6•825294 | 9 | -31•4482 |
| k_12/k_13/k_23/k_34 Interference | Negative Binomial | 81•00482 | 80•8957 | 6•824613 | 10 | -30•4478 |
| k_34 Interference | Poisson | 81•19591 | 81•15432 | 7•083238 | 6 | -34•5772 |
| k_13/k_14/k_23/k_34 Interference | Negative Binomial | 81•27301 | 81•16388 | 7•092799 | 10 | -30•5819 |
| k_13/k_23/k_24/k_34 Interference | Negative Binomial | 81•33726 | 81•22813 | 7•157046 | 10 | -30•6141 |
| Independent | Poisson | 81•34561 | 81•31592 | 7•244836 | 5 | -35•658 |
| k_12/k_14/k_23/k_24/k_34 Interference | Poisson | 81•43387 | 81•32474 | 7•253661 | 10 | -30•6624 |
| k_23/k_34 Interference | Negative Binomial | 81•49641 | 81•42505 | 7•35397 | 8 | -32•7125 |
| k_12/k_13/k_23/k_24/k_34 Interference | Poisson | 81•49798 | 81•38885 | 7•317765 | 10 | -30•6944 |
| k_14/k_23/k_24/k_34 Interference | Negative Binomial | 81•50433 | 81•3952 | 7•324116 | 10 | -30•6976 |
| k_12/k_13/k_14/k_24 Interference | Negative Binomial | 81•62264 | 81•51351 | 7•442426 | 10 | -30•7568 |
| k_14/k_23 Interference | Poisson | 81•65501 | 81•59954 | 7•528454 | 7 | -33•7998 |
| k_12/k_13/k_24 Interference | Negative Binomial | 81•66243 | 81•57319 | 7•502106 | 9 | -31•7866 |
| k_12/k_14/k_34 Interference | Negative Binomial | 81•73941 | 81•65017 | 7•579084 | 9 | -31•8251 |
| k_12 Interference | Poisson | 81•79849 | 81•7569 | 7•68582 | 6 | -34•8785 |
| k_13/k_14/k_23/k_24/k_34 Interference | Negative Binomial | 81•83156 | 81•70055 | 7•629462 | 11 | -29•8503 |
| k_12/k_14/k_23/k_34 Interference | Poisson | 81•95657 | 81•86733 | 7•796249 | 9 | -31•9337 |
| k_12/k_24/k_34 Interference | Poisson | 82•00625 | 81•93489 | 7•863811 | 8 | -32•9674 |
| k_12/k_23/k_34 Interference | Poisson | 82•02646 | 81•9551 | 7•884021 | 8 | -32•9776 |
| k_12/k_13/k_14/k_23/k_34 Interference | Negative Binomial | 82•03961 | 81•9086 | 7•837514 | 11 | -29•9543 |
| k_14/k_34 Interference | Poisson | 82•0537 | 81•99823 | 7•927147 | 7 | -33•9991 |
| Complete Interference | Poisson | 82•06698 | 81•93596 | 7•864876 | 11 | -29•968 |
| k_14/k_24/k_34 Interference | Negative Binomial | 82•44348 | 82•35424 | 8•283157 | 9 | -32•1771 |
| k_12/k_14 Interference | Poisson | 82•44702 | 82•39155 | 8•320466 | 7 | -34•1958 |
| k_12/k_13/k_14/k_24/k_34 Interference | Negative Binomial | 82•51493 | 82•38392 | 8•312832 | 11 | -30•192 |
| k_14 Interference | Poisson | 82•55851 | 82•51692 | 8•44584 | 6 | -35•2585 |
| k_12/k_23/k_24/k_34 Interference | Poisson | 82•97089 | 82•88165 | 8•810566 | 9 | -32•4408 |
| k_13/k_23/k_34 Interference | Negative Binomial | 82•99501 | 82•90577 | 8•834683 | 9 | -32•4529 |
| k_13/k_24/k_34 Interference | Poisson | 83•06446 | 82•99311 | 8•922024 | 8 | -33•4966 |
| k_12/k_13/k_23/k_24 Interference | Negative Binomial | 83•10911 | 82•99998 | 8•928897 | 10 | -31•5 |
| k_34 Interference | Negative Binomial | 83•2221 | 83•16663 | 9•095547 | 7 | -34•5833 |
| Independent | Negative Binomial | 83•29309 | 83•25151 | 9•180423 | 6 | -35•6258 |
| k_12/k_13/k_23/k_24/k_34 Interference | Negative Binomial | 83•42906 | 83•29804 | 9•226957 | 11 | -30•649 |
| k_12/k_34 Interference | Negative Binomial | 83•62575 | 83•55439 | 9•483307 | 8 | -33•7772 |
| k_12/k_13/k_14/k_23/k_24 Interference | Negative Binomial | 83•65427 | 83•52325 | 9•452165 | 11 | -30•7616 |
| k_12 Interference | Negative Binomial | 83•79043 | 83•73495 | 9•663871 | 7 | -34•8675 |
| k_12/k_14/k_24/k_34 Interference | Poisson | 83•79888 | 83•70963 | 9•638551 | 9 | -32•8548 |
| k_14/k_34 Interference | Negative Binomial | 83•93824 | 83•86688 | 9•795795 | 8 | -33•9334 |
| k_14/k_23 Interference | Negative Binomial | 83•93944 | 83•86808 | 9•797 | 8 | -33•934 |
| k_23 Interference | Negative Binomial | 83•94815 | 83•89268 | 9•821597 | 7 | -34•9463 |
| Complete Interference | Negative Binomial | 84•00843 | 83•85352 | 9•782433 | 12 | -29•9268 |
| k_12/k_23 Interference | Negative Binomial | 84•12068 | 84•04932 | 9•978237 | 8 | -34•0247 |
| k_12/k_14 Interference | Negative Binomial | 84•35198 | 84•28062 | 10•20954 | 8 | -34•1403 |
| k_14/k_24 Interference | Poisson | 84•50737 | 84•4519 | 10•38081 | 7 | -35•2259 |
| k_14/k_23/k_34 Interference | Negative Binomial | 85•07472 | 84•98548 | 10•91439 | 9 | -33•4927 |
| k_14/k_23/k_24 Interference | Poisson | 85•75936 | 85•688 | 11•61692 | 8 | -34•844 |
| k_12/k_14/k_24/k_34 Interference | Negative Binomial | 85•7671 | 85•65797 | 11•58689 | 10 | -32•829 |
| k_12/k_14/k_24 Interference | Poisson | 85•9602 | 85•88884 | 11•81775 | 8 | -34•9444 |
| k_12/k_23 Interference | Poisson | 86•12684 | 86•07137 | 12•00029 | 7 | -36•0357 |
| k_14/k_24 Interference | Negative Binomial | 86•37207 | 86•30071 | 12•22962 | 8 | -35•1504 |
| k_12/k_34 Interference | Poisson | 86•52753 | 86•47205 | 12•40097 | 7 | -36•236 |
| k_14 Interference | Negative Binomial | 86•59552 | 86•54005 | 12•46897 | 7 | -36•27 |
| k_12/k_14/k_23 Interference | Negative Binomial | 86•64693 | 86•55769 | 12•48661 | 9 | -34•2788 |
| k_12/k_14/k_23/k_24/k_34 Interference | Negative Binomial | 87•03761 | 86•90659 | 12•8355 | 11 | -32•4533 |
| k_12/k_14/k_23/k_24 Interference | Poisson | 87•37491 | 87•28567 | 13•21459 | 9 | -34•6428 |
| k_14/k_23/k_24 Interference | Negative Binomial | 87•61352 | 87•52428 | 13•4532 | 9 | -34•7621 |
| k_12/k_14/k_23/k_34 Interference | Negative Binomial | 87•6937 | 87•58457 | 13•51349 | 10 | -33•7923 |
| k_12/k_14/k_24 Interference | Negative Binomial | 87•84414 | 87•7549 | 13•68381 | 9 | -34•8774 |
| k_12/k_23/k_24 Interference | Poisson | 87•86622 | 87•79486 | 13•72378 | 8 | -35•8974 |
| k_12/k_14/k_23 Interference | Poisson | 88•30729 | 88•23593 | 14•16485 | 8 | -36•118 |
| k_12/k_14/k_23/k_24 Interference | Negative Binomial | 88•99551 | 88•88638 | 14•8153 | 10 | -34•4432 |
| k_12/k_23/k_34 Interference | Negative Binomial | 89•43365 | 89•34441 | 15•27333 | 9 | -35•6722 |
| k_24/k_34 Interference | Poisson | 89•95594 | 89•90047 | 15•82938 | 7 | -37•9502 |
| k_12/k_24 Interference | Poisson | 90•15407 | 90•0986 | 16•02751 | 7 | -38•0493 |
| k_24 Interference | Poisson | 90•24374 | 90•20215 | 16•13107 | 6 | -39•1011 |
| k_23/k_24 Interference | Poisson | 90•29491 | 90•23943 | 16•16835 | 7 | -38•1197 |
| k_23/k_24/k_34 Interference | Poisson | 91•43645 | 91•36509 | 17•29401 | 8 | -37•6825 |
| k_24/k_34 Interference | Negative Binomial | 92•11024 | 92•03889 | 17•9678 | 8 | -38•0194 |
| k_12/k_24 Interference | Negative Binomial | 92•16258 | 92•09122 | 18•02014 | 8 | -38•0456 |
| k_12/k_24/k_34 Interference | Negative Binomial | 92•22393 | 92•13469 | 18•0636 | 9 | -37•0673 |
| k_24 Interference | Negative Binomial | 92•29386 | 92•23838 | 18•1673 | 7 | -39•1192 |
| k_12/k_23/k_24 Interference | Negative Binomial | 93•33413 | 93•24489 | 19•17381 | 9 | -37•6224 |
| k_23/k_24 Interference | Negative Binomial | 93•41608 | 93•34472 | 19•27364 | 8 | -38•6724 |
| k_23/k_24/k_34 Interference | Negative Binomial | 93•56769 | 93•47845 | 19•40736 | 9 | -37•7392 |
| k_12/k_23/k_24/k_34 Interference | Negative Binomial | 93•68065 | 93•57152 | 19•50044 | 10 | -36•7858 |

## Evidence before this study

We searched PubMed for the publications until November 07, 2019 using search terms: (“Plasmodium” AND “falciparum” OR “ovale” OR “malariae” AND “interactions” AND “prevalence”). We retrieved 121 studies that reported species interaction and prevalence. There were two studies published that investigated longitudinal prevalence of *P. ovale* spp. and *P. malariae* (non-falciparum) malaria in Africa. The study by Roucher et al., was a longitudinal study which investigated non-falciparum malaria in a single village in Senegal from 1990 to 2010. Diagnosis was done using microscopy rather than more sensitive PCR. In this study, the prevalence and burden of non-falciparum decreased dramatically with the introduction of ACTs. Similarly, the study by Yman et al., which also investigated prevalence of non-falciparum malaria, took place in a single village in Tanzania over a 22-year period starting in 1994. This study used PCR for speciation, although they did not distinguish the two *P. ovale* species. This study showed persistent transmission of *P. ovale* and *P. malariae* despite declining *P. falciparum*. Neither study, however, has explored whether between species interactions occur or incorporated statistical modelling to assess for interactions.

## Added value of this study

Our study describes the prevalence of non-falciparum malaria longitudinally in different malaria endemic zones of Kenya, while comprehensively evaluating symptoms and clinical data associated with these infections. This is the first study to investigate longitudinal prevalence of non-falciparum across a range of malaria endemicities, while also speciation between *P. ovale curtisi* and *P. ovale wallikeri*. The addition of *P. ovale* spp. speciation enabled the development of novel statistical models to test for between species interactions, revealing a significant interference between *P. falciparum* and *P. ovale curtisi*. This finding, in combination with the different rates of increase in *P. ovale wallikeri* and *P. ovale curtisi* add to the growing evidence that these two species have different within-host adaptations. In addition, the symptomatic presentation of infection at clinic increases our understanding of the range of symptoms associated with non-falciparum malaria.

### Implications of all the available evidence

Our findings show that about 27.5% of the naturally occurring infections in Kenya contain non-falciparum species. This is significantly higher than previous data based on microscopy based diagnosis of non-falciparum malaria. Further, we have shown that non-falciparum malaria infections have been increasing since the introduction of ACT as first-line treatment in Kenya. This could also explain the recent upsurge documented in travelers’ malaria caused by non-falciparum. We also provide evidence that malaria symptoms such as fever, headache, joint pains and seizure can be caused by non-falciparum infections as single or double infections in the absence of *P. falciparum*. This study demonstrated a shift in the prevalence of non-falciparum malaria, with *P. ovale* spp. infections increasing over time. Viewed together, our findings suggest that non-falciparum infections must be increasingly considered in malaria control efforts if continued reductions in malaria prevalence are to occur.

---

Akala et al., **Characterising *Plasmodium* inter-species interactions during a period of increasing prevalence of *Plasmodium ovale***

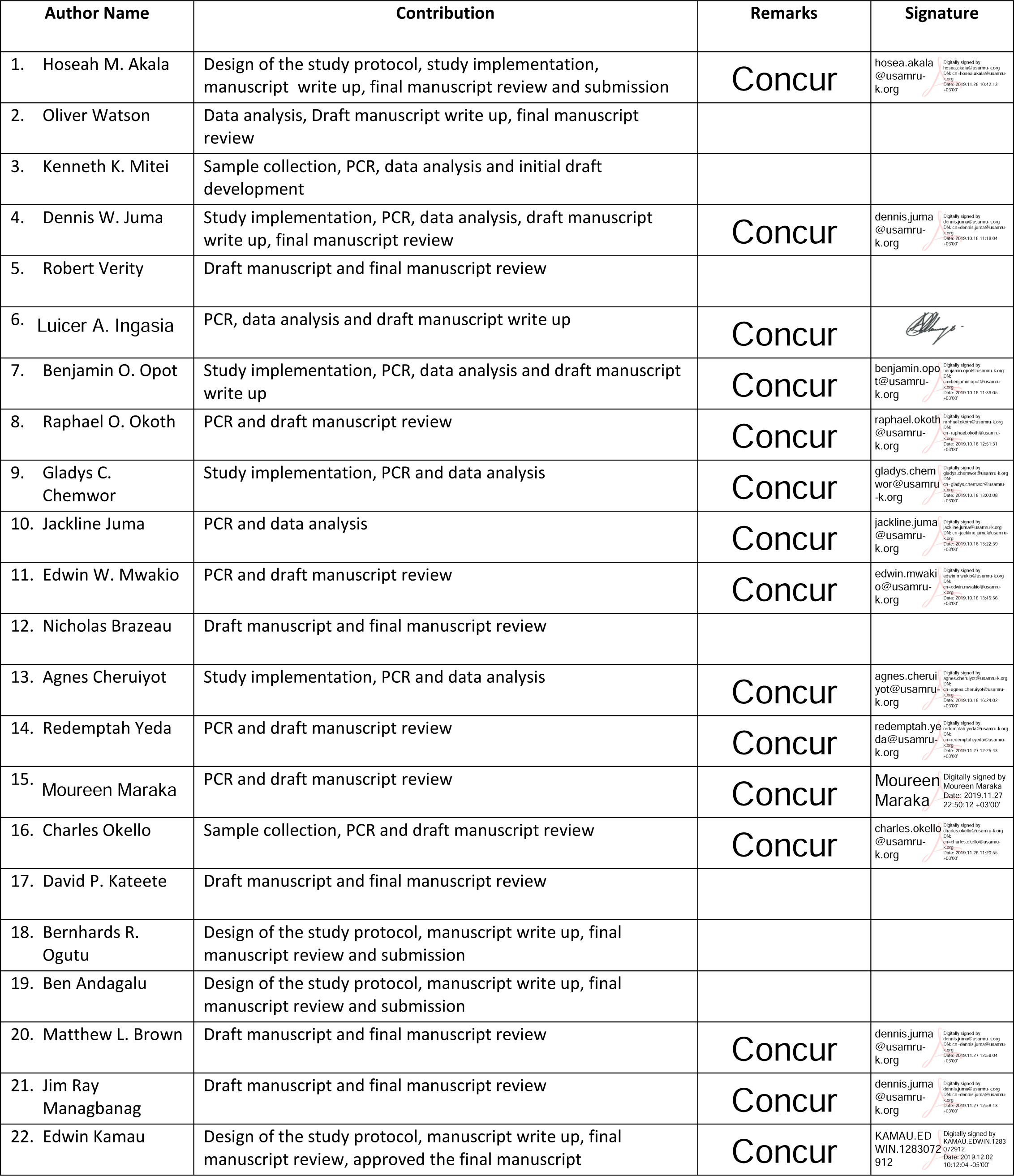
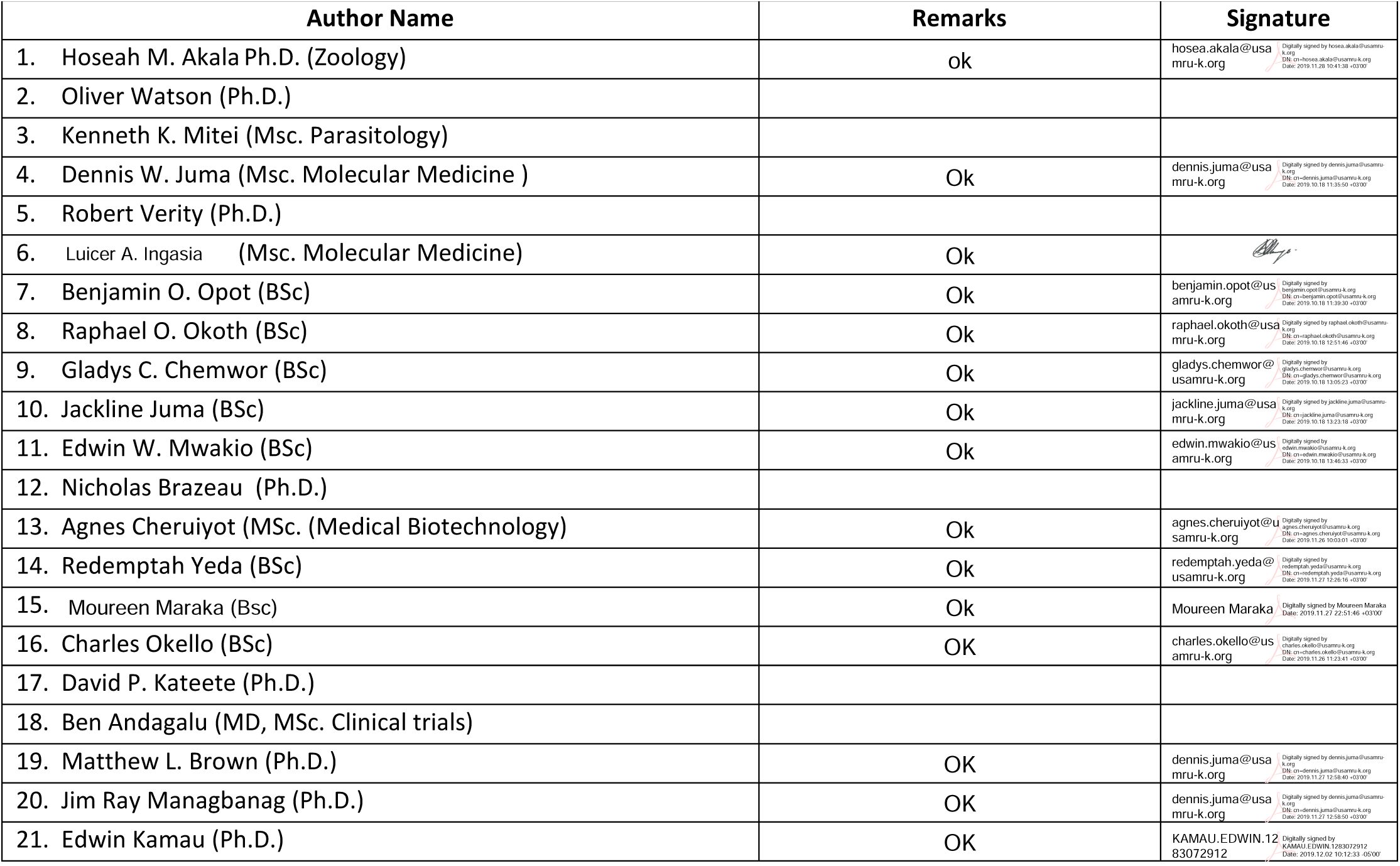

H M Akala PhD, D W Juma MSc, B O Opot BSc, R O Okoth BSc, G C Chemwor BSc, J A Juma BSc, E W Mwakio BSc, A C Cheruiyot MSc, R A Yeda1 BSc, M N Maraka BSc, C O Okello BSc, B Andagalu MSc, J R Managbanag Ph.D.; - Department of Emerging and Infectious Diseases (DEID), United States Army Medical Research Directorate-Africa (USAMRD-A), Kenya Medical Research Institute (KEMRI) / Walter Reed Project, P. O. Box 54 – 40100, Kisumu, Kenya.

Luicer A. Ingasia - University of the Witwatersrand

O Watson Ph.D., R Verity Ph.D.; - Medical Research Council, Centre for Global Infectious Disease Analysis, Department of Infectious Disease Epidemiology, Imperial College London

K K Mitei MSc; - Department of Emerging and Infectious Diseases (DEID), United States Army Medical Research Directorate-Africa (USAMRD-A), Kenya Medical Research Institute (KEMRI) / Walter Reed Project, P. O. Box 54 – 40100, Kisumu, Kenya.^3^College of Health Sciences, Makerere University, Kampala, Uganda,

D P Kateete Ph.D.; - College of Health Sciences, Makerere University, Kampala, Uganda L A Ingasia MSc;

N Brazeau Ph.D.; - Department of Epidemiology, Gillings School of Global Public Health, University of North Carolina, Chapel Hill, NC

M L Brown Ph.D.; - Walter Reed Army Institute of Research 503 Robert Grant Ave, Silver Spring, MD 20910, United States

E Kamau Ph.D.; - U.S. Military HIV Research Program, Walter Reed Army Institute of Research, Silver Spring, MD, 6720A Rockledge Drive, Suite 400, Bethesda, MD 20817, United States of America.

